# Non-additive effects of schizophrenia risk genes reflect convergent downstream function

**DOI:** 10.1101/2023.03.20.23287497

**Authors:** PJ Michael Deans, Carina Seah, Jessica Johnson, Judit Garcia Gonzalez, Kayla Townsley, Evan Cao, Nadine Schrode, Eli Stahl, Paul O’Reilly, Laura M. Huckins, Kristen J. Brennand

## Abstract

Genetic studies of schizophrenia (SCZ) reveal a complex polygenic risk architecture comprised of hundreds of risk variants, the majority of which are common in the population at-large and confer only modest increases in disorder risk. Precisely how genetic variants with individually small predicted effects on gene expression combine to yield substantial clinical impacts in aggregate is unclear. Towards this, we previously reported that the combinatorial perturbation of four SCZ risk genes (“eGenes”, whose expression is regulated by common variants) resulted in gene expression changes that were not predicted by individual perturbations, being most non-additive among genes associated with synaptic function and SCZ risk. Now, across fifteen SCZ eGenes, we demonstrate that non-additive effects are greatest within groups of functionally similar eGenes. Individual eGene perturbations reveal common downstream transcriptomic effects (“convergence”), while combinatorial eGene perturbations result in changes that are smaller than predicted by summing individual eGene effects (“sub-additive effects”). Unexpectedly, these convergent and sub-additive downstream transcriptomic effects overlap and constitute a large proportion of the genome-wide polygenic risk score, suggesting that functional redundancy of eGenes may be a major mechanism underlying non-additivity. Single eGene perturbations likewise fail to predict the magnitude or directionality of cellular phenotypes resulting from combinatorial perturbations. Overall, our results indicate that polygenic risk cannot be extrapolated from experiments testing one risk gene at a time and must instead be empirically measured. By unravelling the interactions between complex risk variants, it may be possible to improve the clinical utility of polygenic risk scores through more powerful prediction of symptom onset, clinical trajectory, and treatment response, or to identify novel targets for therapeutic intervention.

## INTRODUCTION

The genetic architecture of schizophrenia (SCZ) is complex and polygenic, comprised predominantly of common risk variants (342 to date^1^) found in non-coding regions and thought to regulate the expression of one or more genes^2-4^. Their proximal *cis*-targets (termed “eGenes”) are enriched for genes expressed in glutamatergic neurons^5,6^ and with roles in synaptic function and gene regulation^1,7-12^. A long-standing question is how risk variants with individually minuscule effects combine to yield substantial impacts in aggregate (reviewed^13^). Thus, we ask how the effects of eGenes sum, interact, and converge within and across critical biological functions.

We previously applied CRISPR-engineering of human induced pluripotent stem cell (hiPSC)-based models to study four top-ranked SCZ eGenes (*FURIN, SNAP91, TSNARE1, CLCN3*), revealing an unexpected combinatorial effect not predicted from single gene perturbations that was concentrated on synaptic function and SCZ risk^14^. Albeit, three of the four eGenes tested have roles in synaptic biology (*SNAP91*^15^, *TSNARE1*^16^, *CLCN3*^17^). Here we expand upon this finding, evaluating combinatorial interactions between fifteen top SCZ eGenes. We test the hypothesis that non-additive effects are a consequence of shared biological function, contrasting the extent of non-additivity observed when manipulating groups of biologically related genes, specifically those with known roles at the synapse (“synaptic”: *SNAP91, CLCN3, PLCL1, DOC2A, SNCA*), or regulating transcription (“regulatory”: *ZNF823, INO80E, SF3B1, THOC7, GATAD2A*), to those with un-related non-synaptic non-regulatory biology (“multi-function”: *CALN1, CUL9, TMEM219, PCCB, FURIN)*, and random combinations thereof.

Individual perturbation of these fifteen eGenes revealed substantial shared transcriptomic changes with a common direction of effect (“convergence”). Combinatorial eGene perturbations within but not across biological functions resulted in transcriptomic changes that were predominantly smaller than predicted by the additive model (“sub-additive effects”). These sub-additive genes overlapped with the convergent gene targets of the same eGenes, were enriched for SCZ risk, and constituted a large proportion of the genome-wide polygenic risk score. Single eGene perturbations failed to predict the magnitude or directionality of combinatorial effects, which resulted in decreased neurite outgrowth, reduced synaptic proteins, and neuronal hyperactivity. Our findings demonstrate that combinatorial interactions between eGenes^18,19^ must be empirically measured rather than predicted from single eGene studies.

## RESULTS

### Prioritization of synaptic, regulatory, and multi-function brain eGenes regulated by SCZ

Genes implicated in synaptic biology and epigenetic/transcriptional regulation are enriched for the common variants^1,7-12^ associated with risk for SCZ. To generate three groups of top eGenes linked to either synaptic biology, gene regulation, or neither (**Fig. 1**), fifteen eGenes were prioritized from the Psychiatric Genomics Consortium (PGC3)-SCZ genome wide association study (GWAS)^1^, using complementary approaches to integrate GWAS and expression quantitative locus (eQTL) information (**Fig. 1A**). First, colocalization of fine-mapped GWAS^1^ loci with post-mortem brain^20^ eQTLs identified 25 loci with very strong evidence (using the Bayesian test COLOC, defining colocalization based on a high posterior probability that a single shared variant is responsible for both signals, PP4 > 0.8^21^). Second, transcriptomic imputation (using prediXcan)^22-24^ identified ∼250 significant genes (p<6×10^−6^) with brain^20^-specific genetically regulated gene expression (GREX) predicted to be associated with SCZ. There was significant overlap between the two analyses, with 22/25 COLOC genes identified by prediXcan (binomial test p-value 3.03×10^−112^). For each eGene, the magnitude and direction of perturbation associated with SCZ risk was predicted. Expression of each SCZ-eQTL gene was confirmed in our hiPSC neuron RNAseq^14^. eGenes were further separated into discrete functional categories based on gene ontology annotations (http://geneontology.org/). Our final gene list included five synaptic genes (*SNAP91, CLCN3, PLCL1, DOC2A, SNCA*), five regulatory genes (*ZNF823, INO80E, SF3B1, THOC7, GATAD2A*), and five genes with non-synaptic, non-regulatory functions, termed “multi-function” (*CALN1, CUL9, TMEM219, PCCB, FURIN*) (**Table 1**).

**Table 1.** Top SCZ-GWAS eGenes prioritized as synaptic (green), regulatory/epigenetic (blue), and multi-function (purple).

|  |  |  |  |  |  |  |  |  |
| --- | --- | --- | --- | --- | --- | --- | --- | --- |
|  | GWAS |  |  |  |  | COLOC | PrediXcan |  |
| Group | Chr | SNP | P | LD range | Gene | PP4 | z | P |
| synapse | 6 | rs2022265 | 3.74E-10 | 83262613...85419243 | *SNAP91* | 0.893265 | 5.185282 | 2.16E-07 |
| synapse | 4 | rs356183 | 3.37E-08 | 89645368...91759130 | *SNCA* | 0.847582 | -4.494898 | 6.96E-06 |
| synapse | 16 | rs3814883 | 8.82E-15 | 30016830...30034591 | *DOC2A* | 0.521296 | 6.088335 | 1.14E-09 |
| synapse | 4 | rs61405217 | 5.39E-11 | 170533784...170644824 | *CLCN3* | 0.718835 | 5.771783 | 7.84E-09 |
| synapse | 2 | rs1451488 | 6.72E-17 | 198669426...199437305 | *PLCL1* | 0.035058 | 4.910285 | 9.09E-07 |
| regulatory | 19 | rs72986630 | 3.07E-10 | 10832225...12839819 | *ZNF823* | 0.999253 | 6.350252 | 2.15E-10 |
| regulatory | 16 | rs3814883 | 8.82E-15 | 29007489...31016970 | *INO80E* | 0.894347 | 7.639276 | 2.18E-14 |
| regulatory | 3 | rs704364 | 8.41E-10 | 62819885...64848612 | *THOC7* | 0.893413 | -5.236222 | 1.64E-07 |
| regulatory | 2 | rs2914983 | 1.10E-14 | 197256700...199299078 | *SF3B1* | 0.888092 | 7.256793 | 3.96E-13 |
| regulatory | 19 | rs1858999 | 7.97E-14 | 18497024...20619093 | *GATAD2A* | 0.858178 | -7.214675 | 5.41E-13 |
| proprotein convertase | 15 | rs4702 | 2.15E-23 | 90414642...92426654 | *FURIN* | 0.999943 | -10.06869 | 7.60E-24 |
| ubiquitination | 6 | rs113113059 | 2.29E-11 | 42150013...44192158 | *CUL9* | 0.953677 | 5.754229 | 8.70E-09 |
| calcium signalling | 7 | rs2944821 | 1.90E-09 | 70244499...72912040 | *CALN1* | 0.913966 | 5.547481 | 2.90E-08 |
| metabolism | 3 | rs7432375 | 5.32E-15 | 134969406...137055316 | *PCCB* | 0.906413 | -7.547429 | 4.44E-14 |
| signalling | 16 | rs3814883 | 8.82E-15 | 28952638...30984212 | *TMEM219* | 0.846736 | 6.291546 | 3.14E-10 |

**Figure 1.**
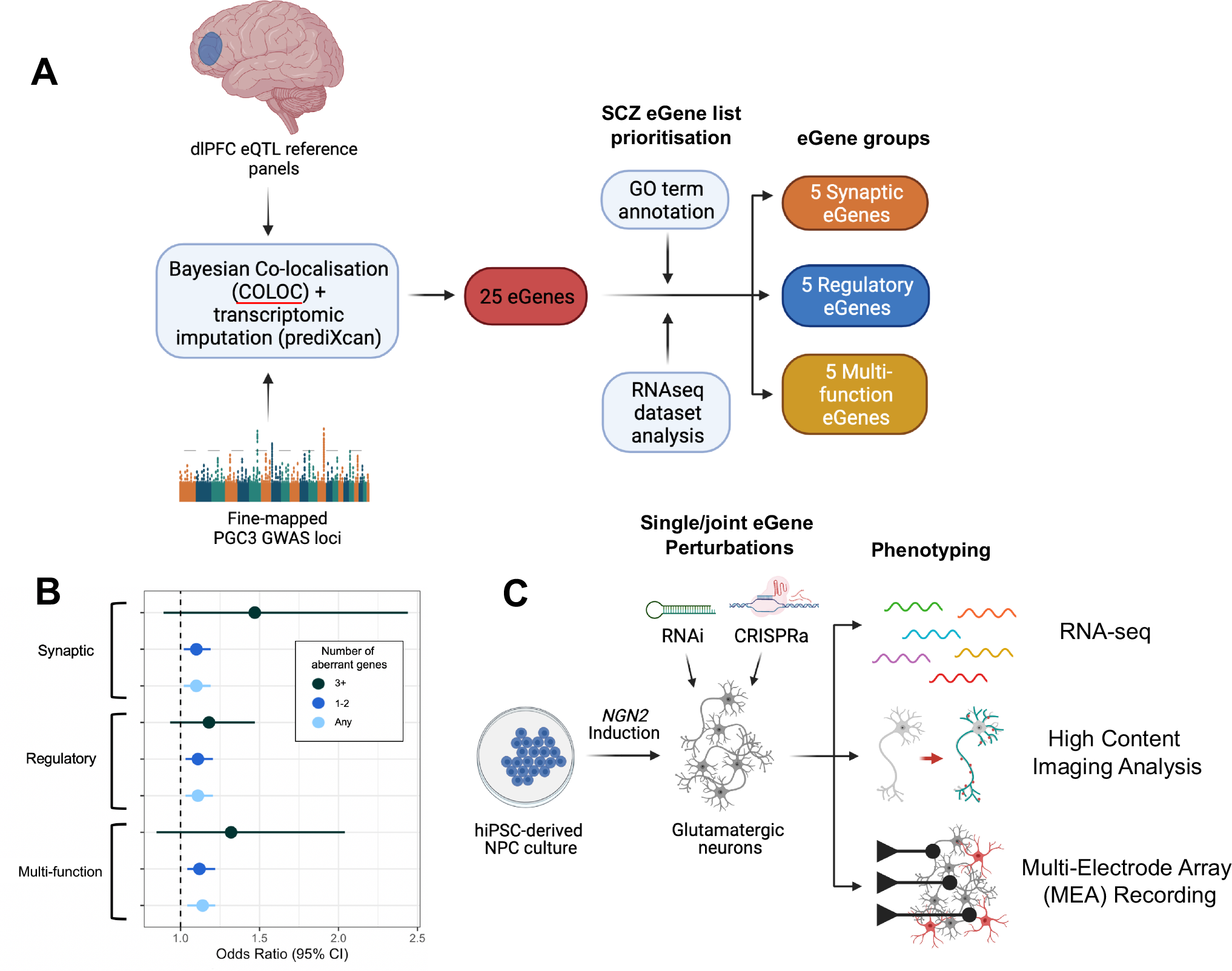
Prioritization and manipulation of synaptic, regulatory, and multi-function brain eGenes regulated by SCZ. **A**. Schematic of SCZ eGene identification and prioritization. Fine-mapped GWAS loci were co-localized with post-mortem brain eQTLs using COLOC, identifying 25 candidate SCZ eGenes. Transcriptomic imputation using prediXcan identified a further ∼250 significant genes brain-specific genetically regulated gene expression (GREX) predicted to be associated with SCZ. 22/25 eGenes identified using COLOC overlapped with the prediXcan gene list. For further study, 15 of these eGenes were separated into three functional groups based on gene ontology annotations. **B**. Predicted GREX levels in dorsolateral prefrontal cortex (DLPFC) calculated for the fifteen eGenes in a Swedish SCZ cohort. Aberrant expression of eGenes is predicted to impact SCZ case-control status in a dose-dependent manner. **C**. Summary schematic of experimental set up interrogating functions of eGenes. Individual and joint perturbation of eGenes using CRISPRa and RNAi in hiPSC-derived glutamatergic neuron cultures was followed by neuronal phenotyping using three modalities: RNA-seq, high content imaging and multi-electrode array recording.

Predicted GREX levels in dorsolateral prefrontal cortex (DLPFC) were calculated for the fifteen eGenes in a Swedish SCZ cohort^25^ (see **Methods**). Aberrant GREX, defined as GREX in the top or bottom deciles of expression, significantly predicted SCZ case-control status (p<0.01774, OR>1.10); with the highest odds ratio when three or more genes were perturbed (OR_3_=1.47 vs. OR_1_=1.10), suggesting a dose-dependent effect (**Fig. 1B**).

### Shared transcriptomic impact of SCZ eGenes related to synaptic function

Transient overexpression of *NGN2* rapidly induces functionally mature human glutamatergic neurons (iGLUTs). We^14,26-28^ and others^29-36^ demonstrated iGLUTs to be >95% glutamatergic neurons, robustly express glutamatergic genes, release neurotransmitters, produce spontaneous synaptic activity, and recapitulate the impact of psychiatric disease associated genes. iGLUTs expressed most SCZ eGenes (70% >0.5 log2RPKM, 63% >1 log2RPKM), including all fifteen eGenes prioritized herein (>2.8 log2RPKM).

To explore the transcriptomic impact of manipulating SCZ eGenes alone and in combination (**Fig. 2, SI Fig. 2**), CRISPRa and shRNAs were used to increase and decrease, respectively, endogenous expression of SCZ eGenes in the direction associated with SCZ risk (**Fig. 1C, SI Fig. 1D**). Three to five vectors per gene were tested in 7-day-old (D7) iGLUTs, in order to select the single vector that best achieved the level of perturbation predicted by eQTL analyses (**SI Fig. 1D**). Each eGene was perturbed in 21-day-old (D21) iGLUTs for 72 hours (**SI Fig. 1E, SI Fig 2A, SI Fig 4A**), individually and jointly, including appropriate vector and scrambled controls, from two neurotypical donors with average polygenic risk scores (one experimental batch per donor). Three groups of five randomly combined genes, one group of ten randomly combined genes, and one group of all fifteen genes were also included.

**Figure 2.**
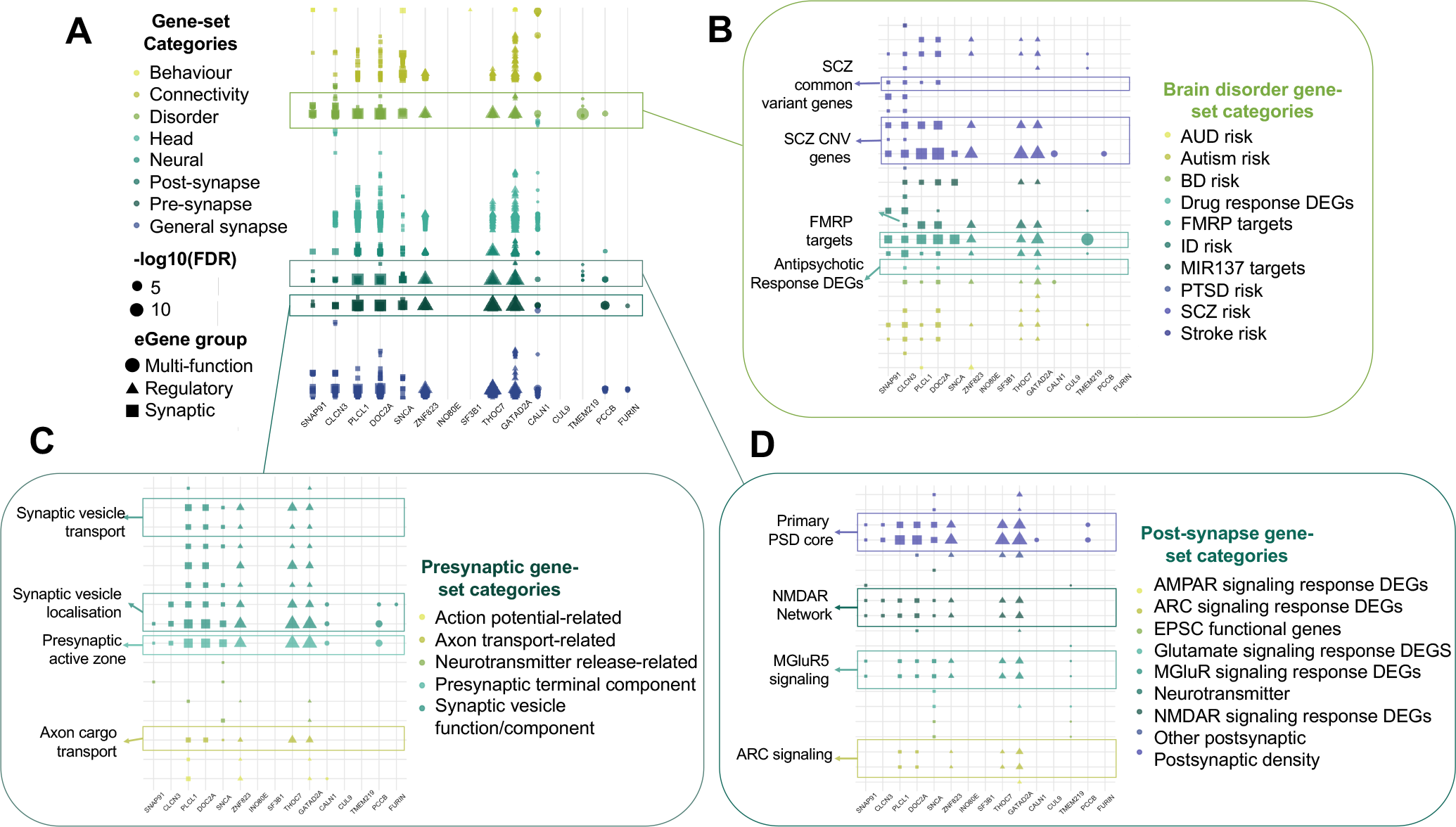
Perturbation of SCZ eGenes results in differential expression of genes relating to brain disorders and synaptic function. **A**. Gene set enrichment analysis (GSEA) performed across a collection of 698 manually curated gene-sets with a neural theme revealed enrichments of gene-sets related to brain disorders and synaptic functions across 12/15 eGene perturbations. **B**. Across brain disorder-related gene-sets, 5/5 Synaptic eGene perturbations showed strong enrichment of other genes relating to schizophrenia risk, including both common variant-linked genes and copy number variant (CNV) genes associated with schizophrenia. C. Across presynaptic function gene-sets, 10/15 eGene perturbations showed enrichment of genes relating to synaptic vesicle localization and transport. **D**. Across postsynaptic function gene-sets, 8/15 eGene perturbations showed enrichment of genes relating to components of glutamatergic neurotransmission. AUD = alcohol use disorder, BD = bipolar disorder, FMRP = Fragile X Mental Retardation Protein, ID = intellectual disability, PTSD = post traumatic stress disorder, SCZ = schizophrenia, DEGs = differentially expressed genes.

For all fifteen eGene perturbations, competitive gene-set enrichment analysis (GSEA) was applied within the differentially expressed genes (DEGs, p_FDR_<0.05) using 698 manually curated neural gene-sets^37^ (**Fig. 2A)**, revealing strong enrichment of presynaptic (**Fig. 2C**), postsynaptic (**Fig. 2D)** and brain-disorder-related (**Fig. 2B)** gene-sets. The majority of eGene perturbations (10/15) resulted in DEGs that were strongly enriched for gene-sets related to synaptic biology and glutamatergic neurotransmission. Enrichments for SCZ GWAS (4/15), SCZ copy number variant (CNV) (10/15), and antipsychotic response (3/15) gene-sets were also frequently observed (**Fig. 2B-D**). Altogether, individual manipulation of fifteen eGenes, towards approximating the magnitude and direction of SCZ risk, resulted in a shared transcriptomic impact related to synaptic biology and SCZ risk.

### Combinatorial perturbation of SCZ eGenes with shared biological functions results in sub-additive impacts on transcription

Differential expression following simultaneous perturbation of all five eGenes within each of the synaptic, regulatory and multi-function groups (“measured combinatorial”) was compared to the sum of differential expression for each single eGene perturbation (“expected additive”) (**Fig. 3; SI Fig. 4-5, Box 1**). The difference between the measured combinatorial and expected additive model for each group represents the observed non-additive effects (**Fig. 3A**). Combinatorial perturbations of synaptic, regulatory, and multi-function eGenes resulted in non-additive effects across 16.8%, 20.2%, and 0% of the total transcriptome (**Fig. 3B**). This was not explained by the magnitude of eGene perturbation between individual and combinatorial perturbations, which did not significantly differ (p>0.05 Wilcoxon ranked sum test). The size of the observed non-additive transcriptional effect also increased directly with the total number of eGenes simultaneously perturbed, indicating that increasing the number of eGenes perturbed may increase the degree of interactive effects on transcription (**SI Fig. 5B**).

**Figure 3.**
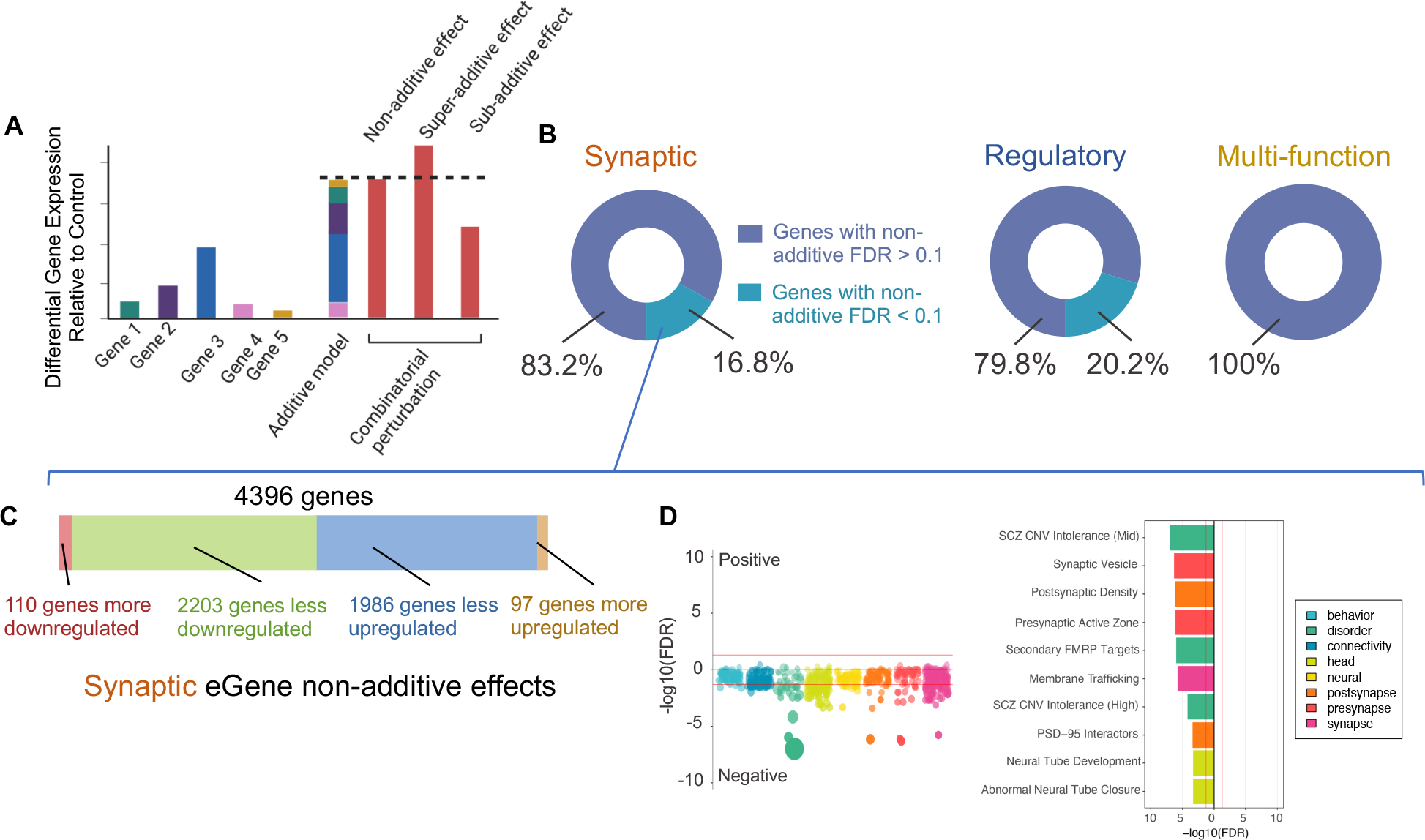
Perturbation of SCZ eGenes within functional categories results in non-additive effects on transcription impacting expression of genes linked to brain disorders and synaptic function. **A**. Schematic of differential expression analysis. Individual eGene perturbations, the implementation of the expected additive model based on the latter and the measured combinatorial perturbation permitting the detection of interactive effects through comparison with the additive model. **B**. Combinatorial perturbation of eGenes within, but not across functional pathways resulted in non-additive effects on expression across 16.8% (Synaptic eGenes) and 20.2% (Regulatory eGenes) of the transcriptome. **C**. Following joint perturbation of the Synaptic eGene set, the majority (>95%) of non-additive genes showed significantly less differential expression in the measured combinatorial perturbation relative to the expected additive model (“less downregulated” and “less upgregulated” categories). **D**. GSEA of non-additive genes in the Synaptic eGene set demonstrated significant enrichment for genes relating to brain disorders and synaptic function. SCZ = schizophrenia, CNV = copy number variant, FMRP = Fragile X Mental Retardation Protein, FDR = false discovery rate. See **Box 1** for example schematics and explanation for each sub-category of non-additive effect.

Unexpectedly, the majority (>95%) of genes exhibiting non-additive effects in the synaptic and regulatory groups demonstrated less differential expression than predicted by the additive model (“sub-additive effects”) (**Fig. 3C, SI Fig. 4B, Box 1**). For both synaptic and regulatory eGenes, this sub-additive set of genes were enriched for synaptic and brain disorder-associated gene-sets (**Fig. 3D, SI Fig. 4C**). Although sub-additive effects might imply a failure to achieve combinatorial perturbations at the single cell level, consistent with the high-titer lentiviral gRNAs used here, we note that for the four SCZ eGenes previously evaluated^14^, observed non-additive effects were similar whether tested from a single vector expressing all gRNAs^14^ or from independent expression vectors (**SI Fig. 3A-F**). Overall, combinatorial perturbation of eGenes within synaptic or regulatory but not multifunction groups resulted in sub-additive perturbations in genes enriched for synaptic biology and SCZ risk.

### Sub-additive transcriptomic effects reflect convergence and co-regulation of downstream targets

To test the hypothesis that sub-additive effects arose from shared downstream signatures of individual eGene perturbations, DEGs were meta-analyzed within each functional group (using METAL^38^, p< 1.92E-06), and “convergent” target genes defined as those with both shared direction of effect across all five eGene perturbations and with non-significant heterogeneity (Cochran’s heterogeneity Q-test p_Het_ > 0.05). (**Fig. 4, SI Fig. 6**). There was robust within-group convergence for the synaptic (1070 genes) and regulatory (1070 genes) eGene groups, but limited convergence across the multi-function eGenes (71 genes) (**Fig. 4B-E**). Most of the convergent genes overlapped with the sub-additive genes (Fisher’s exact test, p<2.2E-16 for both synaptic and regulatory eGene groups) (**Fig. 4C, D**). Across all eight combinations of eGenes studied here (synaptic, regulatory, multi-function and random combinations of five, ten and fifteen eGenes), convergence across individual perturbations is correlated with the degree of non-additive effect seen in the corresponding joint perturbation condition (**Fig 4A**, Pearson’s r^2^ = 0.6569, p=0.0147). Convergent targets of synaptic eGenes were enriched for synaptic function (e.g. pre-synaptic active zone, p=7.45E-04) and brain disorder (e.g. SCZ GWAS, p=1.08E-06) gene-sets (**Fig. 4F-H**). In contrast, convergent targets of regulatory eGene effects were enriched for brain disorder gene-sets only (e.g. bipolar disorder, p=9.92E-06) (**SI Fig. 6)**. Taken together, these findings highlight converging effects of SCZ eGenes on specific targets relating to synaptic function and brain disorders.

**Figure 4.**
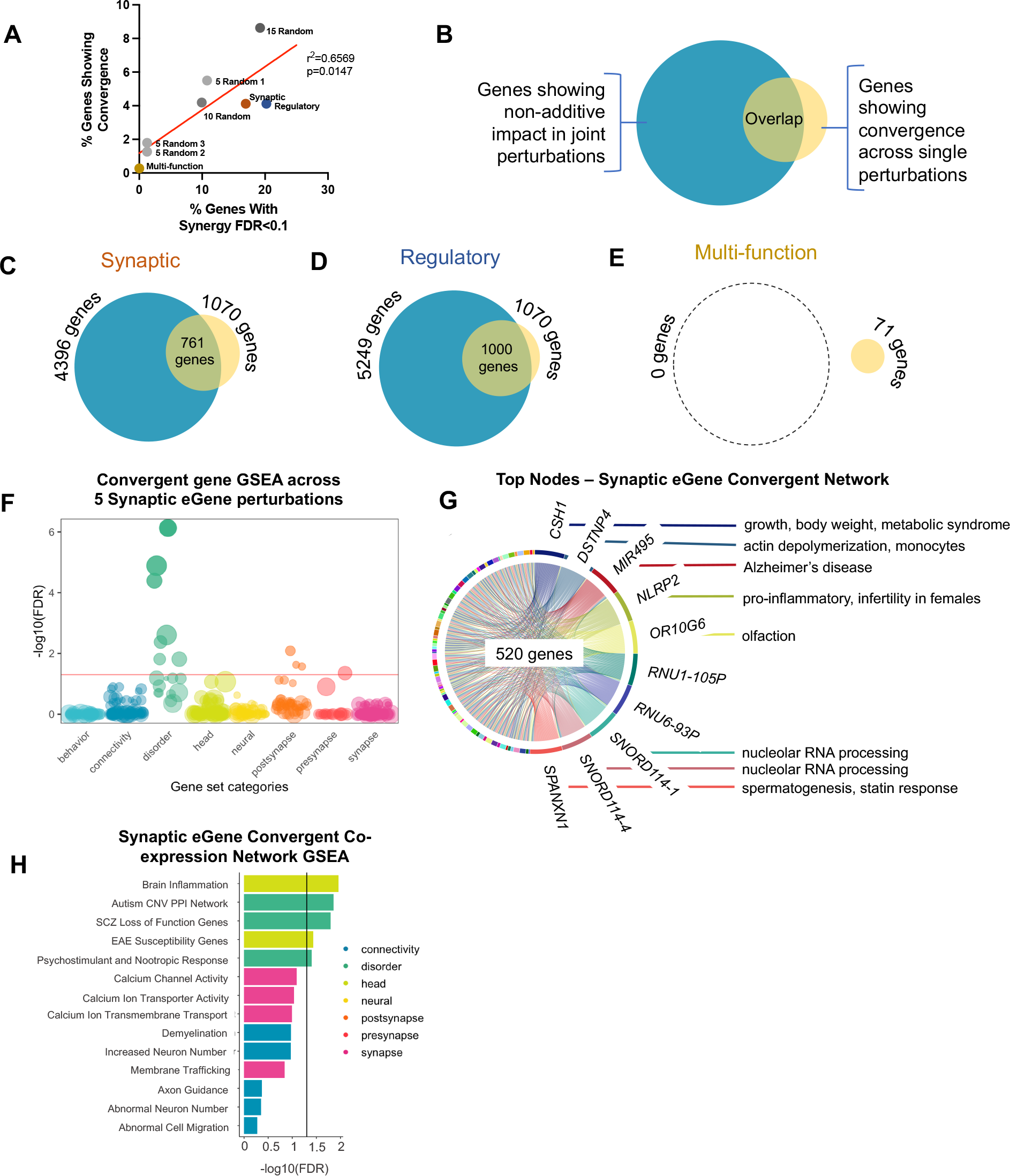
Convergence accounts for non-additive effects within functional pathways. **A-E**. Meta-analysis of differentially expressed genes (DEGs) elicited by individual eGene perturbations for each five-gene grouping using METAL to identify DEGs that showed altered expression consistently in the same direction across all five eGene perturbation conditions for each set of eGenes. **A**. Convergence across individual eGene perturbations is correlated with the degree of non-additive effect seen in the corresponding joint perturbation condition. Pearson’s r^2^ = 0.6569, p=0.0147. **B**. For each joint eGene perturbation group, non-additive impacts on transcription were compared with genes showing significant convergence across individual perturbations for the same eGene set. **C**. Evidence of convergence was found in 1070 genes across the synaptic eGene perturbations, 761 of which also exhibited non-additive effects in the additive-combinatorial comparison for the same set. **D**. Evidence of convergence was found in 1070 genes across the regulatory eGene perturbations, 1000 of which also exhibited non-additive effects in the additive-combinatorial comparison for the same set. **E**. No significant non-additive effects and only minimal convergence could be seen in eGene perturbations across functional pathways. **F**. GSEA of convergent genes in the Synaptic eGene set demonstrated significant enrichment for genes relating to brain disorders and presynaptic functions. **G**. Bayesian biclustering identified significant convergence of co-expression networks unique to synaptic pathways that replicated in over 25% of iterations. Major node genes mediating convergent networks of synaptic eGenes included *OR10G6*, an olfactory receptor gene, and *MIR495*, which has previously been implicated in Alzheimer’s disease. **H**. GSEA of co-expressed network genes in the Synaptic eGene set demonstrated significant enrichment for genes relating to brain inflammation and schizophrenia risk. GSEA = gene-set enrichment analysis; SCZ = schizophrenia; CNV = copy number variant; PPI = protein-protein interaction; EAE = Experimental Autoimmune Encephalomyelitis; FDR = false discovery rate.

To mechanistically probe the co-regulatory structure underlying downstream convergence of single gene effects, we next assessed co-regulated networks. We performed Bayesian bi-clustering and network reconstruction across within-function groups. We performed unsupervised bi-clustering and reconstruction over 40 runs to identify high-confidence co-expressed gene networks that replicated across a maximum number of iterations. From this, we identified co-regulated networks across synaptic and regulatory eGene perturbations. Significant convergence of co-expression networks unique to synaptic pathways replicated in over 25% of iterations, while shared regulatory networks replicated at a lower confidence of 12.5%. Networks shared across synaptic eGene perturbations comprised of 520 genes enriched for inflammatory pathways and SCZ risk (**Fig. 4G-H**), while the regulatory eGene downstream network was wider, encompassing 676 genes enriched for SCZ risk, calcium signaling, and synaptic function (**SI Fig. 6B, C**). This wider network with less replication across perturbations is consistent with the broad functional coverage of eGenes within this pathway. The smaller number of synaptic eGene subnetworks replicating at higher confidence suggests tighter co-regulation, corresponding to the higher degree of sub-additive effect observed. Likewise, the multi-function eGene group did not exhibit significant convergent networks, consistent with the lack of non-additive effects observed with combinatorial CRISPR-perturbation (**Fig. 4E)**.

Altogether, the overlap of sub-additive genes resulting from combinatorial eGene perturbations with the convergent targets downstream of individual eGene manipulations suggest that eGenes with shared biological functions impact common downstream networks. This redundancy suggests that saturation of directional impacts on gene expression might represent the mechanism underlying non-additive effects.

### Non-additive genes are associated with SCZ risk

PRS are not yet sufficiently accurate for individual-level clinical utility^39,40^, and most genome-wide PRSs are presently calculated in a way that distills an individual’s genetic liability to a single number, ignoring biological function^41^. Here, we explored the non-additive signatures in the context of GWAS and polygenic prediction of case-control SCZ status, to validate the relevance and specificity of our findings to SCZ risk and pathophysiology (**Fig. 5; SI Fig. 7**). Specifically, we tested (1) whether non-additive genes are enriched in schizophrenia GWAS signal, and (2) to what extent non-additive genes are part of the total genetic risk load to schizophrenia. Pathway PRS for non-additive genes from each functional category were calculated using PRSet^42^ (see **Methods**), and results were compared with standard, genome-wide PRS (PRSice)^43^ and with pathway PRS from curated lists of 1233 synaptic genes (Synapse Gene Ontology^44^) and 1639 transcription factors (The Human Transcription Factor database^45^).

**Figure 5.**
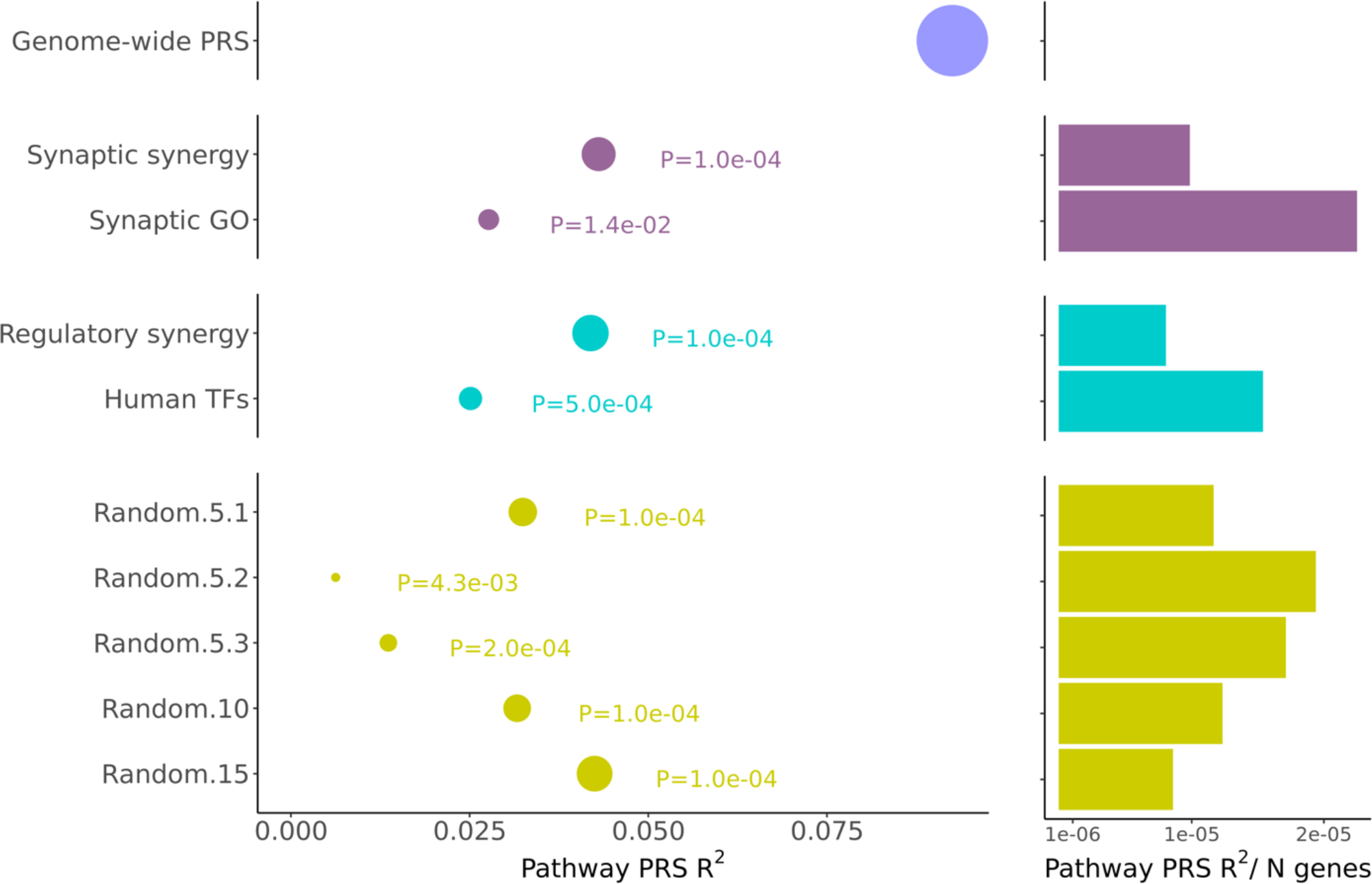
Within and across-pathway synergistic genes are associated with SCZ risk. Left: phenotypic variance explained (R^2^) by genome-wide (purple), synaptic (magenta), regulatory (blue) and random (green) pathway PRS. The size of the dot represents number of genes included in each pathway/gene set. The P next to each dot represents the empirical “competitive” *P*-value calculated by PRSet to evaluate pathway enrichment. Right: phenotypic variance explained (R^2^) normalized by the number of genes within each pathway.

All non-additive-specific PRSs (derived from 4,834 SCZ cases and 6,128 controls from the Swedish population-based cohort^46^) showed significant association with SCZ case/control status and enrichment in SCZ GWAS signal, regardless of the number of SNPs/genes included in the PRS (**Fig. 5A**, left panel). Pathway PRSs composed of non-additive genes in relation to synaptic (4,306 genes in PRS; R^2^=0.0431), regulatory (5,249 genes in PRS; R^2^=0.0419) and all fifteen eGenes (4,988 genes in PRS, R^2^=0.0425) explained the largest proportion of variance in SCZ case-control status, accounting for almost half of the total genome-wide PRS (19,340 genes plus SNPs in regions outside gene annotations in genome-wide PRS, R^2^= 0.0925). PRSet enrichment results from the Synaptic Gene Ontology (**SI Fig. 7A-B**) were in line with previous reports from the SCZ PGC-3 GWAS^1^, highlighting post-synaptic ontology terms (**SI Fig. 7C**).

The strength of the PRSs was proportional to the number of genes included in the pathway; after correcting by the number of genes included (**Fig. 5A**, right panel) and comparing same-sized pathways (**SI Fig. 7D**), non-additive genetic signal did not differ between groups. Overall, these results show that non-additive genes comprise a large part of the total genome-wide genetic risk, regardless of non-additive genes being obtained from within or across functional groups. Our results also suggest that –although non-additive genes are enriched in GWAS signal in aggregation– individually they do not harbor greater polygenic signal than curated synaptic genes.

### Individual gene perturbations do not predict combinatorial effects at the cellular level

Finally, the consequences of combinatorial perturbations on neuronal morphology, synaptic density, and neuronal activity were evaluated, towards resolving whether the interactions between individual eGene effects seen at the level of gene expression were reflected in cellular phenotypes (**Fig. 1C, Fig, 6, SI Fig. 1E**, **SI Fig. 8-9**).

High content imaging of D7 iGLUTs assessed neurite outgrowth and D21 iGLUTs assessed synaptic puncta (SYN1+) density relative to total length of MAP2AB+ dendrites. Many individual eGene perturbations did not yield detectable impacts on neurite outgrowth or synaptic puncta density; notable exceptions were that perturbation of *FURIN*, which was associated with a significant decrease in neurite outgrowth, consistent with our previous findings^14^, and individual perturbation of all regulatory eGenes resulted in significantly increased puncta density. Intriguingly, combinatorial perturbation of synaptic eGenes led to significant reductions in both neurite outgrowth (**Fig. 6A**,**D, SI Fig. 8A)** and puncta density (**Fig. 6B**,**D, SI Fig. 8B)**, in a manner opposite to the increases predicted by the expected additive model. Likewise, combinatorial perturbation of regulatory eGenes led to a significant reduction in puncta density, again opposite to the prediction of the expected additive model (**Fig. 6D, SI Fig. 8B)**. Simultaneous perturbation of all five multi-function genes did not result in any significant impact on either neurite outgrowth or puncta density (**Fig. 6D, SI Fig. 8B)**.

**Figure 6.**
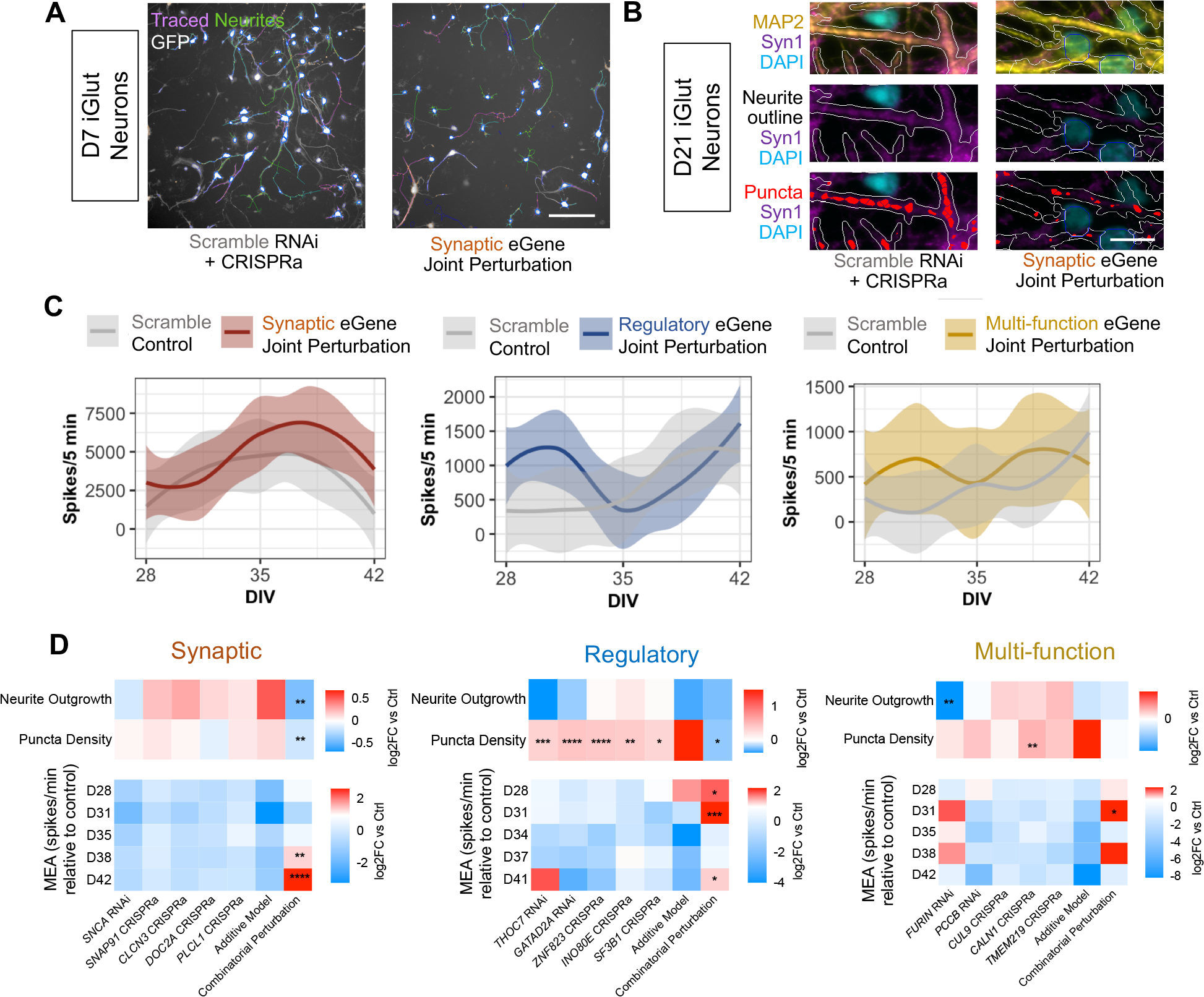
Combinatorial perturbation of SCZ eGenes within functional categories results in impaired neurite outgrowth, synaptic expression and neuronal hyperactivity. **A**. Combinatorial perturbation of all five synaptic eGenes in D7 iGLUTs resulted in significant reduction of neurite outgrowth relative to a combinatorial scramble gRNA + shRNA control. **B**. Combinatorial perturbation of all five synaptic eGenes in D21 iGLUTs resulted in significant reduction of Syn1+ puncta density relative to a combinatorial scramble gRNA + shRNA control. **C**. LOESS plots; combinatorial perturbation of eGenes within but not across functional categories results in transient neuronal hyperactivity in MEA recordings of D28-42 iGLUTs. **D**. Summary heatmap of neurite outgrowth, puncta density and neuronal activity data for within and across function eGene perturbations. N = minimum of 2 independent experiments across 2 donor lines with 10-12 technical replicates per condition. One-way ANOVA with post-hoc Bonferonni multiple comparisons test. * = p<0.05; ** = p<0.01; *** = p<0.001; **** = p<0.0001.

Axion multi-electrode arrays (MEA) measured firing frequency, burst firing patterns and coordinated network firing in iGLUTs over time. Here again, individual perturbations of eGenes did not typically result in detectable impacts on neuronal firing frequency. Combinatorial perturbation of the synaptic or regulatory eGenes was associated with transient neuronal hyperactivity within two weeks post perturbation, a pattern not predicted by the individual eGene perturbations (**Fig. 6C, D, SI Fig. 9**).

Unexpectedly, the sub-additive effects observed at the transcriptomic level manifested instead as phenotypic differences at the cellular level that are distinct and opposite from those predicted by individual eGene perturbations.

## DISCUSSION

Perturbations of SCZ eGenes resulted in non-additive and convergent transcriptional effects that occur within groups of functionally similar genes (see also ^47^) and increased with number of eGenes perturbed. Across fifteen eGenes, within-function combinatorial perturbations resulted in sub-additive impacts on transcriptional profiles of hundreds of genes, which largely overlapped with the convergent gene targets of the same SCZ eGenes. Thus, at the transcriptional level, sub-additive effects likely reflected a saturation of the impact at convergent downstream targets. This provides clear evidence, for the first time in relation to SCZ risk, for functional redundancy in biological function-specific perturbations. Our observations demonstrate that polygenic risk cannot be dissected by extrapolating from experiments that test one risk gene at a time; combinatorial interactions should be empirically measured as they are poorly modelled using current frameworks. This unexpected finding may apply more broadly across complex genetic disorders.

In contrast, at the cellular level, non-additive effects typically manifested as a phenotypic change in the opposite direction to what was predicted by individual eGene perturbations. At this time, we cannot resolve the extent to which these opposing combinatorial effects at the phenotypic level reflect compensatory homeostatic responses not predicted by gene expression, differences in the timing and duration of combinatorial eGene perturbations during neurodevelopment across the transcriptomic and phenotypic experimental paradigms, and/or the possibility of ameliorative non-cell-autonomous effects of the human astrocyte co-culture used for phenotypic assays. Most critically, transcriptomic and phenotypic impacts of combinatorial perturbations reflect emergent properties of neuronal biology that are not well-predicted by single eGene perturbations alone. In addition to expanding the number of eGenes and biological functions evaluated, future studies should manipulate SCZ eGenes across more neuronal and glia cell types and developmental stages, investigating how cell-type-specific expression patterns and network co-expression modules^26^ impact additivity, interactivity and convergence. As we systematically explore the hundreds of eGenes linked to psychiatric disorders, the importance of non-additive effects in disease risk will become clearer.

The robust overlap between non-additive genes and convergent gene targets suggest a functional redundancy of within-function eGenes, suggesting that the observed sub-additive impact reflected a saturation of the downstream effects of individual eGenes converging on common gene targets. Using a pooled CRISPR screen of a partially overlapping group of 10 SCZ eGenes in iGLUTs, we similarly reported that the strength of convergence increased with gene number and that the specificity of convergence increased with the degree of functional similarity and co-expression between eGenes^47^. Convergent genes downstream of SCZ eGenes were showed cross-disorder enrichment targets for psychiatric risk genes, suggesting that they may underlie shared clinical hallmarks and pleiotropy of risk^47^. Moreover, recent studies of loss-of-function autism spectrum disorder genes, albeit perturbations unlikely to be inherited in combination, also reported convergence, whether evaluated *in vitro* in human neural progenitor cells^48^ and brain organoids^49^, or *in vivo* in fetal mouse brains^50^ and *Xenopus tropicalis*^51^. Further exploration of cross-disorder convergence of risk is certainly warranted: the common and rare risk variants for SCZ^1,7-12,52-54^, autism spectrum disorder^55-57^ and more broadly across the neuropsychiatric disorder spectrum^58-62^ are all highly enriched for genes involved in synaptic biology and gene regulation. Altogether, while individual CRISPR-mediated genetic perturbation of SCZ eGenes in iGLUTs reveal their causal impacts on gene expression, neurodevelopment, and neuronal activity^32,63-65^, sub-additive and convergent effects suggests a model to explain non-additive interactions and argue for the prioritization of convergent genes as potential therapeutic targets.

Additive and non-additive impacts remain difficult to assess using genomic approaches. Recall that for the fifteen eGenes, aberrant DLPFC GREX significantly predicted SCZ case-control status in a dose-dependent manner (**Fig. 1B**). To test whether the aggregate effect of these eGenes was also additive, Transcriptomic Risk Scores (TRS) were calculated (see **Methods**). In general, TRS constructed from larger (p<2.2 × 10^−16^) (**SI Fig. 1B**) or more biologically diverse (R=0.19, p<2.2×10^−16^) (**SI Fig. 1C**) gene groups more significantly predicted SCZ. Overall, extreme predicted expression differences of SCZ eGenes predict SCZ case-control status in a dose-dependent but not function-specific manner. Thus, while we show that cell-based within-individual non-additive impacts increase with extent of polygenicity and within groups of functionally similar genes, population-level SCZ risk increases with the number of genes and pathways impacted. This distinction may reflect the lack of individuals, either case or control, with strong within-function perturbations imputed or measured. Conversely, it is also possible that predicted gene expression of proximal target genes does not comprehensively capture the multi-omic impact of perturbing eGenes observed empirically. In either case, we posit that at present, hiPSC-based models are uniquely well suited for testing the *cis*- and *trans*-effects of SCZ risk that cannot yet be resolved through existed genetic and genomic approaches.

While our results are largely consistent with the polygenic additive model^66^, our findings also suggest that a fraction of risk variants may have non-additive effects to a degree dependent on the extent to which their target genes converge within the same biological roles^67^. Others have also reported evidence of compound effects of multiple genes on synaptic development and function in iGLUTs^68^. Moving forward, it is critical to resolve the extent and context within which risk variants sum linearly^18,19^ or are amplified^69^/buffered^70^ by epistatic interactions, towards improving the predictive power and clinical utility of genetic risk scores.

Our data argues for a re-conceptualization of genetic contribution to disease risk; PRSs that incorporate biological pathways and gene co-expression networks of the risk variants being summed^42,71^, together with their downstream impacts^47^, may improve patient stratification^42,72^ or better predict drug response^73^. Moreover, resolving convergent effects shared between eGenes will identify nodes whereby the impact of aggregate genetic risk can be most effectively decreased and prioritize potential therapeutic targets. We seek to move towards precision medicine^74^, whereby the identity and interactions between each patient’s genetic variants inform diagnosis, prognosis, and treatment.

## MATERIALS AND METHODS

### SCZ eGene Prioritization

Colocalization (using COLOC^21,75^) identified 25 fine-mapped GWAS (65,205 cases and 87,919 controls) and eQTL (537 EUR samples)^20^ associations with very strong evidence (PP4>0.8) of colocalization (using COLOC^21,75^). Transcriptomic imputation (using PrediXcan^24^) identified ∼250 genes with significant (p<6×10^−6^)^24^ predicted differential expression between SCZ-cases and controls using PGC3-SCZ GWAS^1^ and post-mortem CommonMind Consortium (CMC)^20^ data (623 samples). Of the 25 COLOC genes, 22 were also significant by PrediXcan. From these 22, we prioritized the top genes across three broad categories: synaptic, regulatory, and multifunction (defined as not synaptic, regulatory, and seemingly unrelated to each other). To complete selection of five genes from each category, three additional top-ranked synaptic genes from the prediXcan analysis were included: *DOC2A*^76^, *CLCN3*^76^ and *PLCL1*. Prioritized genes were not located in the major histocompatibility complex (MHC) locus and had confirmed expression in iGLUTs.

### NGN2-glutamatergic neuron (iGLUT) induction from hiPSC-derived NPCs^28,33^

Validated control hiPSCs for CRISPRa/RNAi were selected from a previously reported case/control hiPSC cohort of childhood onset SCZ (COS)^77^. The following controls^77^ were used: hiPSC NPCs NSB553-S1-1 (male), NSB2607-1-4 (male).

hiPSC-NPCs expressing dCas9-VPR were generated and validated as previously described^77^ and cultured in hNPC media (DMEM/F12 (Life Technologies #10565), 1x N2 (Life Technologies #17502-048), 1x B27-RA (Life Technologies #12587-010), 1x Antibiotic-Antimycotic, 20 ng/ml FGF2 (Life Technologies)) on Matrigel (Corning, #354230). hiPSC-NPCs at full confluence (1-1.5×10^7^ cells / well of a 6-well plate) were dissociated with Accutase (Innovative Cell Technologies) for 5 mins, spun down (5 mins X 1000g), resuspended and seeded onto Matrigel-coated plates at 3-5×10^6^ cells / well. Media was replaced every two-to-three days for up to seven days until next split.

At day -2, dCas9-VPR hiPSC-NPCs were seeded as 1.2×10^^6^ cells / well in a 12-well plate coated with Matrigel. At day -1, cells were transduced with rtTA (Addgene 20342) and NGN2 (Addgene 99378) lentiviruses. Medium was switched to non-viral medium four hours post infection. At D0, 1 μg/ml dox was added to induce *NGN2*-expression. At D1, transduced hiPSC-NPCs were treated with corresponding antibiotics to the lentiviruses (300 ng/ml puromycin for dCas9-effectors-Puro, 1 mg/ml G-418 for NGN2-Neo) in order to increase the purity of transduced hiPSC-NPCs. At D3, NPC medium was switched to neuronal medium (Brainphys (Stemcell Technologies, #05790), 1x N2 (Life Technologies #17502-048), 1x B27-RA (Life Technologies #12587-010), 1 μg/ml Natural Mouse Laminin (Life Technologies), 20 ng/ml BDNF (Peprotech #450-02), 20 ng/ml GDNF (Peprotech #450-10), 500 μg/ml Dibutyryl cyclic-AMP (Sigma #D0627), 200 nM L-ascorbic acid (Sigma #A0278)) including 1 μg/ml Dox. 50% of the medium was replaced with fresh neuronal medium once every second day. At D13, iGLUTs were treated with 200 nM Ara-C to reduce the proliferation of non-neuronal cells in the culture, followed by half medium changes. At D18, Ara-C was completely withdrawn by full medium change while adding media containing individual shRNA/gRNA vectors or pools of mixed shRNA and gRNA vectors (Addgene 99374), either targeting eGenes or scramble controls. Medium was switched to non-viral medium four hours post infection. At D19, transduced iGLUTs were treated with corresponding antibiotics to the gRNA lentiviruses (1 mg/ml HygroB for lentiguide-Hygro/lentiguide-Hygro-mTagBFP2) followed by half medium changes until the neurons were harvested for RNA-seq experiments at D21 (Supplemental figure 1E).

### CRISPRa/shRNA Validation^14^

At day -2, dCas9-VPR hiPSC-NPCs were seeded as 0.6×10^^6^ cells / well in a 24-well plate coated with Matrigel. At day -1, cells were transduced with rtTA (Addgene 20342) and NGN2 (Addgene 99378) lentiviruses. Medium was switched to non-viral medium four hours post infection. At D0, 1 μg/ml dox was added to induce *NGN2*-expression. At D1, transduced hiPSC-NPCs were treated with corresponding antibiotics to the lentiviruses (1 mg/ml G-418 for NGN2-Neo) in order to increase the purity of transduced hiPSC-NPCs. At D3, NPC medium was switched to neuronal medium (Brainphys (Stemcell Technologies, #05790), 1x N2 (Life Technologies #17502-048), 1x B27-RA (Life Technologies #12587-010), 1 μg/ml Natural Mouse Laminin (Life Technologies), 20 ng/ml BDNF (Peprotech #450-02), 20 ng/ml GDNF (Peptrotech #450-10), 500 μg/ml Dibutyryl cyclic-AMP (Sigma #D0627), 200 nM L-ascorbic acid (Sigma #A0278)) including 1 μg/ml Dox. 50% of the medium was replaced with fresh neuronal medium once every second day. At D4 individual shRNA/gRNA vectors (Addgene 99374), either targeting eGenes or scramble controls. 3-5 vectors were tested per eGene. Medium was switched to non-viral medium four hours post infection. At D5, transduced iGLUTs were treated with corresponding antibiotics to the gRNA lentiviruses (1 mg/ml HygroB for lentiguide-Hygro/lentiguide-Hygro-mTagBFP2) before harvesting at D7 in order to assess eGene perturbation efficacy via qPCR.

#### Real time-quantitative PCR

Real time qPCR was performed as previously described ^78^. Specifically, cell cultures were harvested with Trizol and total RNA extraction was carried out following the manufacturer’s instructions. Quantitative transcript analysis was performed using a QuantStudio 7 Flex Real-Time PCR System with the Power SYBR Green RNA-to-Ct Real-Time qPCR Kit (all Thermo Fisher Scientific). Total RNA template (25 ng per reaction) was added to the PCR mix, including primers listed below. qPCR conditions were as follows; 48°C for 15 min, 95°C for 10 min followed by 45 cycles (95°C for 15 s, 60°C for 60 s). All qPCR data is collected from at least three independent biological replicates of one experiment. A one-way ANOVA with posthoc Dunnett’s multiple comparisons test was performed on data for the set of targeting vectors for each eGene relative to the scramble control vector. Data analysis were performed using GraphPad PRISM 6 software.

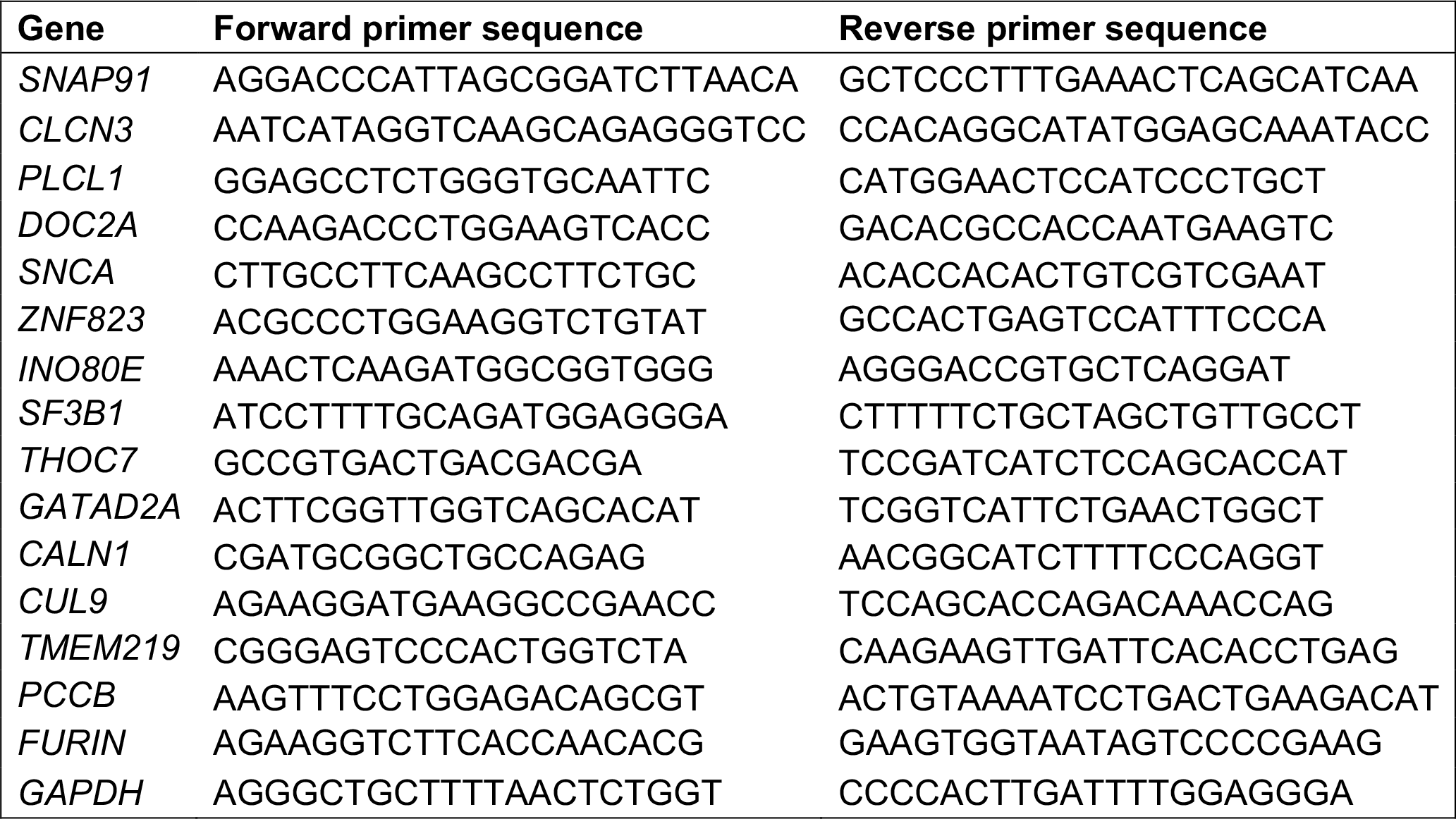

#### Immunostaining and high-content imaging microscopy

##### Neurite analysis

iGLUTs were seeded as 1.5×10^4^ cells/well in a 96-well plate coated with 4x Matrigel at day 3. iGLUTs were plated in media containing individual shRNA/gRNA vectors or pools of mixed shRNA and gRNA vectors (Addgene 99374), either targeting eGenes or scramble controls. Medium was switched to non-viral medium four hours post infection. At day 4, transduced iGLUTs were treated with corresponding antibiotics to the gRNA lentiviruses (1 mg/ml HygroB for lentiguide-Hygro/lentiguide-Hygro-mTagBFP2) followed by half medium changes until the neurons were fixed at day 7 (Supplemental figure 1E). At day 7, cultures were fixed using 4% formaldehyde/sucrose in PBS with Ca^2+^ and Mg^2+^ for 10 minutes at room temperature (RT). Fixed cultures were washed twice in PBS and permeabilised and blocked using 0.1% Triton/2% Normal Donkey Serum (NDS) in PBS for two hours. Cultures were then incubated with primary antibody solution (1:1000 MAP2 anti chicken (Abcam, ab5392) in PBS with 2% NDS) overnight at 4°C. Cultures were then washed 3x with PBS and incubated with secondary antibody solution (1:500 donkey anti chicken Alexa 647 (Life technologies, A10042) in PBS with 2% NDS) for 1 hour at RT. Cultures were washed a further 3x with PBS with the second wash containing 1 μg/ml DAPI. Fixed cultures were then imaged on a CellInsight CX7 HCS Platform with a 20x objective (0.4 NA) and neurite tracing analysis performed using the neurite tracing module in the Thermo Scientific HCS Studio 4.0 Cell Analysis Software. 12-24 wells were imaged per condition across a minimum of 2 independent cell lines, with 9 images acquired per well for neurite tracing analysis. A one-way ANOVA with a post hoc Bonferroni multiple comparisons test was performed on data for neurite length per neuron using Graphpad Prism.

##### Synapse analyses

Commercially available primary human astrocytes (pHAs, Sciencell, #1800; isolated from fetal female brain) were seeded on D3 at 0.85×10^4^ cells per well on a 4x Matrigel-coated 96 W plate in neuronal media supplemented with 2% fetal bovine serum (FBS). iGLUTs were seeded as 1.5×10^5^ cells/well in a 96-well plate coated with 4x Matrigel at day 5. Half changes of neuronal media were performed twice a week until fixation. At day 13, iGLUTs were treated with 200 nM Ara-C to reduce the proliferation of non-neuronal cells in the culture. At day 18, Ara-C was completely withdrawn by full medium change while adding media containing individual shRNA/gRNA vectors or pools of mixed shRNA and gRNA vectors (Addgene 99374), either targeting eGenes or scramble controls. Medium was switched to non-viral medium four hours post infection. At day 19, transduced iGLUTs were treated with corresponding antibiotics to the gRNA lentiviruses (1 mg/ml HygroB for lentiguide-Hygro/lentiguide-Hygro-mTagBFP2) followed by half medium changes until the neurons were fixed at day 21 (Supplemental figure 1E). At day 21, cultures were fixed and immunostained as described previously, with an additional antibody stain for Synapsin1 (primary antibody: 1:500 Synapsin1 anti mouse (Synaptic Systems, 106 011); secondary antibody: donkey anti mouse Alexa 568 (Life technologies A10037)). Stained cultures were imaged and analysed as above using the synaptogenesis module in the Thermo Scientific HCS Studio 4.0 Cell Analysis Software to determine SYN1+ puncta number, area and intensity per neurite length in each image. 20 wells were imaged per condition across a minimum of 2 independent cell lines, with 9 images acquired per well for synaptic puncta analysis. A one-way ANOVA with a post hoc Bonferroni multiple comparisons test was performed on data for puncta number per neurite length using Graphpad Prism.

| Antibody | Species | Supplier | Product number | Dilution |
| --- | --- | --- | --- | --- |
| MAP2 | Ck | Abcam | ab5392 | 1:500 |
| SYNAPSIN1 | Ms | Synaptic Systems | 106 011 | 1:500 |
| Alexa 568 anti-Mouse | Ms | Life technologies | A10037 | 1:500 |
| Alexa 647 anti-Chicken | Ck | Life technologies | A10042 | 1:500 |

#### Multiple Electrode array (MEA)

Commercially available primary human astrocytes (pHAs, Sciencell, #1800; isolated from fetal female brain) were seeded on D3 at 1.7×10^4^ cells per well on a 4x Matrigel-coated 48 W MEA plate (catalog no. M768-tMEA-48W; Axion Biosystems) in neuronal media supplemented with 2% fetal bovine serum (FBS). At D5, iGLUTs were detached, spun down and seeded on the pHA cultures at 1.5×10^5^ cells per well. Half changes of neuronal media supplemented with 2% FBS were performed twice a week until day 42. At day 13, co-cultures were treated with 200 nM Ara-C to reduce the proliferation of non-neuronal cells in the culture. At Day 18, Ara-C was completely withdrawn by full medium change. At day 25, a full media change was performed to add media containing individual shRNA/gRNA vectors or pools of mixed shRNA and gRNA vectors (Addgene 99374), either targeting eGenes or scramble controls. Medium was switched to non-viral medium four hours post infection. Electrical activity of iGLUTs was recorded at 37°C twice every week from day 28 to day 42 using the Axion Maestro MEA reader (Axion Biosystems) (Supplemental figure 1E). Recording was performed via AxiS 2.4. Batch mode/statistic compiler tool was run following the final recording. Quantitative analysis of the recording was exported as Microsoft excel sheet. Data from 6-12 biological replicates were analyzed using GraphPad PRISM 6 software or R.

#### RNAseq

RNA Sequencing libraries were prepared using the Kapa Total RNA library prep kit. Paired-end sequencing reads (100bp) were generated on a NovaSeq platform. Raw reads were aligned to hg19 using STAR aligner^79^ (v2.5.2a) and gene-level expression were quantified by featureCounts^80^ (v1.6.3) based on Ensembl GRCh37.70 annotation model. Genes with over 10 counts per million (CPM) in at least four samples were retained. After filtering, the raw read counts were normalized by the voom^81^ function in limma and differential expression was computed by the moderated t-test implemented in limma^82^. Differential gene expression analysis was performed between each CRISPRa/shRNA target group and scramble control group. Bayes shrinkage (limma::eBayes) estimated modified t- and p-values and identified differentially expressed genes (DEGs) based on an FDR <= 0.05 (limma::TopTable)^83^. GO/pathways were evaluated using Gene-set Enrichment Analysis (GSEA)^84^. In these analyses, the t-test statistics from the differential expression contrast were used to rank genes in the GSEA using the R package ClusterProfiler^85^. Permutations (up to 100,000 times) were used to assess the GSEA enrichment P value.

### Analysis of additive and non-additive effects

We applied our published approach to resolve distinct additive and non-additive transcriptomic effects after combinatorial manipulation of genetic variants and/or chemical perturbagens, developed^14^, applied^26^, and described in detail^37^. The expected additive effect was modeled through addition of the individual comparisons; the non-additive effect was modeled by subtraction of the additive effect from the combinatorial perturbation comparison. Fitting of this model for differential expression identifies genes that show a difference in the expected differential expression computed for the additive model compared to the observed combinatorial perturbation. Briefly, the non-additive effect between eGenes was identified using a limma’s linear model analysis. The coefficients, standard deviations and correlation matrix were calculated, using *contrasts.fit*, in terms of the comparisons of interest. Empirical Bayes moderation was applied using the *eBayes* function to obtain more precise estimates of gene-wise variability. P-values were adjusted for multiple hypotheses testing using false discovery rate (FDR) estimation, and differentially expressed genes were determined as those with FDR ≤ 5%, unless stated otherwise.

However, interpretation of the resulting DEGs depends on several factors, such as the direction of fold change (FC) in all three models. To identify genes whose magnitude of change is larger in the combinatorial perturbation vs. the additive model, we categorized all genes by the direction of their change in both models and their log_2_(FC) in the non-additive model. First, log_2_(FC) standard errors (SE) were calculated for all samples. Genes were then grouped into ‘positive non-addition’ if their FC was larger than SE and ‘negative non-addition’ if smaller than -SE. If the corresponding additive model log_2_(FC) showed the same or no direction, the gene was classified as “more” differentially expressed in the combinatorial perturbation than predicted. GSEA was performed on a curated subset of the MAGMA collection using the limma package camera function, which tests if genes are ranked highly in comparison to other genes in terms of differential expression, while accounting for inter-gene correlation. Due to the small sample size in this study and moderate fold changes in some eGene perturbations, changes in gene expression may be small and distributed across many genes. However, powerful enrichment analyses in the limma package may be used to evaluate enrichment based on genes that are not necessarily genome-wide significant, and identify sets of genes for which the distribution of t-statistics differs from expectation. Over-representation analysis (ORA) was performed when subsets of DEGs were of interest; genes of interests were ranked by –log10 (p-value) and enrichment was performed against a background of all expressed genes using the WebGestaltR package.

### Meta-analysis of gene expression across perturbations

We performed a meta-analysis and Cochran’s heterogeneity Q-test (METAL^38^) using the p-values and direction of effects (t-statistic), weighted according to sample size across all sets of perturbations (Target vs. Scramble DEGs). Genes were defined as convergent if they (1) had the same direction of effect across all 5, 10, or 15 target combinations, (2) were Bonferroni significant in our meta-analysis (Bonferroni adjusted p-value <= 0.05), and (3) had a heterogeneity p-value = >0.05.

### Bayesian Bi-clustering to identify Target-Convergent Networks

Target-Convergent gene co-expression Networks (TCN) were built using an unsupervised Bayesian bi-clustering model, BicMix on the log2CPM expression data from all the replicates across each of the 5-target sets and Scramble gRNA jointly^86^. 40 runs of BicMix were conducted on these data and the output from iteration 300 of the variational Expectation-Maximization algorithm was used. The hyperparameters for BicMix were set based on previous extensive simulation studies^87^. Target-Convergent Network reconstruction^88^ was performed to reconstruct convergent networks from BicMix data. Network connections that did not replicate in more than 10% of the runs were excluded and nodes with less than 5 edges were removed from GSEA. Using FUMAGWAS: GENE2FUNC, the protein-coding genes were functionally annotated and overrepresentation gene-set analysis for each network gene-set was performed^89^.

### Dataset for population-level analysis of synergy

Individuals from the Sweden-SCZ Population-Based cohort were obtained from the database of Genotypes and Phenotypes, Study Accession: phs000473.v2.p2 (N_Cases_ = 5,232, N_Controls_ = 6,468)^25^.

### Pathway polygenic risk scores

Pathway specific PRS analyses were performed using PRSice-2 (v2.3.5) on individual genotype data for the Sweden-SCZ Population based cohort. A total of 4,834 individuals diagnosed with SCZ and 6,128 controls were included after quality control. To calculate the scores, we used a version of the summary statistics from the PGC SCZ GWAS that excludes the Sweden-SCZ data to prevent inflation of results. SNPs were annotated to genes and pathways based on GTF files obtained from ENSEMBL (GRCh37.75). To include potential gene regulatory elements, gene coordinates were extended 35 kilobases (kb) upstream and 10 kb downstream of each gene. We excluded from analyses the Major histocompatibility complex region (chr6:25Mb-34Mb), ambiguous SNPs (A/T and G/C) and SNPs not present in both GWAS summary statistics and genotype data.

To obtain empirical “competitive” *P-*values, that assess GWAS signal enrichment while accounting for pathway size, we performed the following permutation procedure: first, a “background” pathway containing all genic SNPs is constructed, and clumping is performed within this pathway. For each pathway with *m* SNPs, *N=*10,000 null pathways are generated by randomly selecting *m* SNPs from the “background” pathway. The competitive *P*-value can then be calculated as

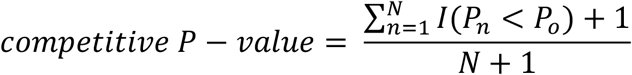

where *I*(.) is an indicator function, taking a value of 1 if the association *P*-value of the observed pathway (*P*_*0*_) is larger than the one obtained from the *n*^th^ null pathway (*P*_*n*_), and 0 otherwise (see^90^ for additional details).

For the analyses testing whether non-additive genes from synaptic/regulatory pathways explain larger R^2^ than *<u>the same number</u>* of non-additive genes from random combinations (**SI Fig. 7**), we took 2,799 random genes from the non-additive synaptic and regulatory transcriptome, which corresponds to the number of genes with non-additive effects in one of the random joint perturbations. For the GTF NULL permutation analyses, we selected n=2,799 random genes from the GTF file GRCh37.75. Pathway PRS for each sample of 2,799 genes was calculated using PRSet^90^, as described above. This procedure was repeated 1000 times.

#### Transcriptomic Risk Score (TRS) Analyses

In order to test the impact of non-additive genetic effects *in silico*, we used transcriptomic imputation methods to calculate genetically-regulated gene expression (GREX) for individuals from the Sweden-SCZ Population-Based cohort. Brain GREX was calculated using PrediXcan^24^ with CMC dorsolateral prefrontal cortex (CMC-DLPFC) models^20^. Predicted GREX levels were calculated for the fifteen eGenes. An initial test of aberrant gene expression was performed by counting the number of genes with dysregulated GREX (defined as predicted GREX in the top or bottom decile of overall expression of that gene, defined in the direction of effect of that gene’s association with SCZ from S-PrediXcan analyses (top decile for positive effect, bottom decile for negative effect) for each of the five-gene groups (synaptic, regulatory, multi-function), and summed the number of aberrant genes present in each individual for each perturbed gene group (Synaptic, Regulatory, and Multi-function). We then looked at the SCZ case/control proportion within each group of individuals with 3+, 1-2, and Any genes with aberrant GREX.

### Association of Synaptic, Regulatory, and Multi-function gene-sets with SCZ

We tested for association of each of the fifteen eGene GREX individually with SCZ (SCZ ∼ GREX), and then calculated composite scores of group GREX (Synaptic, Regulatory, and Multi-function) using a Transcriptomic Risk Score (TRS), calculated as the sum of each GREX weighted by the direction of gene perturbation (1 for activation, -1 for inhibition) from *in vivo* experiments, divided by the total number of genes (N) in the gene-set (Equation X):

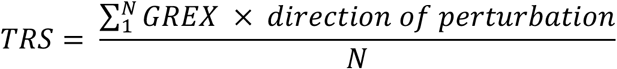

We then tested for association of each TRS (Synaptic, Regulatory, and Multi-function) with SCZ status in the Swedish cohort.

### Permutation tests

We performed permutation tests to assess the impact of (1) the number of genes included in our TRS gene group and (2) the number of pathways impacted by those genes on SCZ case status. We used S-PrediXcan to find genes with CMC-DLPFC GREX associated with SCZ in a large SCZ cohort (N_Cases_ = 11,260, N_Controls_ = 24,542)^8^. From this resulting list of genes, we assigned genes to two groups: nominally-significant genes (N=1,963, Bonferroni p<0.05), and tissue-specific significant genes (N=144, p<0.05/N_Genes in CMC-DLPFC PrediXcan model_). We created pathway sets affected by these genes using the overlap with Kyoto encyclopedia of genes and genomes (KEGG)^91^ and gene ontology (GO)^92,93^. This gave us a sampling pool of 1,465 genes affecting 8,324 pathway sets for the nominally-significant group, and 110 genes affecting 2,382 pathway sets for the tissue-specific group. We then performed permutation sampling analyses (for nominally-significant and tissue-specific significant gene-pathway set pools) where we randomly sampled sets of five, ten, or fifteen genes from the sampling pool (adjusted for the size of each pathway set), calculated TRS from the sampled gene-set, and looked at the association of TRS with SCZ. We performed sampling 100,000 times for each gene-set size. For this analysis, TRS was calculated by taking the sum of each gene in the gene-sets GREX weighted by the direction of effect of the gene association with SCZ from our S-PrediXcan analysis (1 or -1) (Equation X):

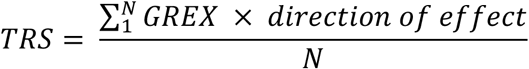

We then looked at the overall association the number of pathways hit by each TRS (based on the annotated lists) with SCZ variance explained (**SI Figure 1A-C**). To determine if the type of pathways hit by our perturbed genes was important to SCZ risk (i.e. is it more important to hit multiple, similar pathways or more diverse pathways to increase SCZ variance explained), we additionally assessed whether the similarity in make-up of pathways affected by the TRS was associated with SCZ. To do this, we used the R “GeneOverlap” package to calculate the average Jaccard Index of pathways for each TRS, and looked at the association of that index with SCZ.

## Data Availability

All source donor hiPSCs have been deposited at the Rutgers University Cell and DNA Repository (study 160; http://www.nimhstemcells.org/); dCas9-VPR hiPSCs are in the process of being submitted in advance of publication.

Processed data and accompanying code can be provided through upon request.

## STATEMENT OF ETHICS

Yale University Institutional Review Board waived ethical approval for this work. Ethical approval was not required because the hiPSC lines, lacking association with any identifying information and widely accessible from a public repository, are thus not considered to be human subjects research. Post-mortem DLPFC data are similarly lacking identifiable information and are not considered human subjects research.

## AUTHOR CONTRIBUTIONS

SCZ eGene lists were prioritized by E.S., L.H., together with K.J.B. *NGN2*-neuronal transcriptomic and phenotypic studies were conducted by P.J.M.D, with assistance by E.C. Synergy analysis was conceptualized by N.S. and applied by P.J.M.D; convergent analyses were conceptualized by K.G.T. and applied by C.S. Transcriptomic imputation was conducted by J.J. and L.H.; pathway-specific PRS by J.G.G. and P.O. The paper was written by P.J.M.D. and K.J.B., with input from all authors.

Special thanks to Michael Talkowski and Douglas Ruderfer for countless discussions on convergence.

## DATA AND CODE AVAILABILITY

Processed data and accompanying code can be provided through upon request.

