## Supplemental Information for "Non-additive effects of schizophrenia risk genes reflect convergent downstream function"

#### SUPPLEMENTAL TABLE

**SI Table 1.** SCZ eGene selection. Highlighted in bold are eGenes prioritized as synaptic (green), regulatory/epigenetic (blue), and unrelated multi-function (purple). (Grey indicates not significant by PrediXcan.)

| | GWAS | | | | | COLOC | PrediXcan | |
|---|---|---|---|---|---|---|---|---|
| Function | Chr | SNP | P | LD range | Gene | PP4 | z | P |
| proprotein convertase | 15 | rs4702 | 2.15E-23 | 90414642…92426654 | ***FURIN*** | 0.999943 | -10.06869 | 7.60E-24 |
| regulatory | 19 | rs72986630 | 3.07E-10 | 10832225…12839819 | ***ZNF823*** | 0.999253 | 6.350252 | 2.15E-10 |
| signalling | 1 | rs56335113 | 3.15E-15 | 28563084…30653243 | *PTPRU* | 0.985465 | -6.850981 | 7.33E-12 |
| metabolism | 8 | rs10957321 | 4.18E-10 | 64500933…66710532 | *CYP7B1* | 0.978623 | -6.202272 | 5.57E-10 |
| development | 17 | rs6504163 | 1.87E-09 | 60555375…62597717 | *ACE* | 0.967474 | -5.869283 | 4.38E-09 |
| pseudogene | 6 | rs2153960 | 9.22E-10 | 108103765…110108063 | *ZNF259P1* | 0.961803 | -5.466066 | 4.60E-08 |
| ubiquitination | 6 | rs113113059 | 2.29E-11 | 42150013…44192158 | ***CUL9*** | 0.953677 | 5.754229 | 8.70E-09 |
| calcium signalling | 7 | rs2944821 | 1.90E-09 | 70244499…72912040 | ***CALN1*** | 0.913966 | 5.547481 | 2.90E-08 |
| signalling | 14 | rs722637 | 9.64E-15 | 103201875…105313861 | *PPP1R13B* | 0.913695 | -5.132138 | 2.86E-07 |
| signalling | 13 | rs9545047 | 3.05E-11 | 79057053…81130204 | *NDFIP2* | 0.907569 | -6.536768 | 6.29E-11 |
| metabolism | 3 | rs7432375 | 5.32E-15 | 134969406…137055316 | ***PCCB*** | 0.906413 | -7.547429 | 4.44E-14 |
| chromatin remodeling | 16 | rs3814883 | 8.82E-15 | 29007489…31016970 | ***INO80E*** | 0.894347 | 7.639276 | 2.18E-14 |
| RNA transport | 3 | rs704364 | 8.41E-10 | 62819885…64848612 | ***THOC7*** | 0.893413 | -5.236222 | 1.64E-07 |
| synapse | 6 | rs2022265 | 3.74E-10 | 83262613…85419243 | ***SNAP91*** | 0.893265 | 5.185282 | 2.16E-07 |
| protein binding | 1 | rs61828917 | 7.95E-10 | 172580537…174637937 | *ANKRD45* | 0.891423 | 3.614873 | 0.0003 |
| splicing | 2 | rs2914983 | 1.10E-14 | 197256700…199299078 | ***SF3B1*** | 0.888092 | 7.256793 | 3.96E-13 |
| signalling | 3 | rs75968099 | 5.16E-11 | 35754053…37781278 | *DCLK3* | 0.88304 | -4.628441 | 3.68E-06 |
| DNA repair | 3 | rs75968099 | 5.16E-11 | 36034966…38107017 | *MLH1* | 0.877109 | 6.376857 | 1.81E-10 |
| metabolism | 11 | rs58950470 | 1.10E-08 | 64482546…66486143 | *RNASEH2C* | 0.869117 | -5.203533 | 1.96E-07 |
| regulatory | 1 | rs11121172 | 7.15E-10 | 7412645…9876635 | *RERE* | 0.867634 | 2.078589 | 0.037655 |
| regulatory | 19 | rs1858999 | 7.97E-14 | 18497024…20619093 | ***GATAD2A*** | 0.858178 | -7.214675 | 5.41E-13 |
| synapse | 4 | rs356183 | 3.37E-08 | 89645368…91759130 | ***SNCA*** | 0.847582 | -4.494898 | 6.96E-06 |
| signalling | 16 | rs3814883 | 8.82E-15 | 28952638…30984212 | ***TMEM219*** | 0.846736 | 6.291546 | 3.14E-10 |
| pseudogene | 2 | rs6546857 | 2.80E-09 | 72872890…74911073 | *ALMS1P* | 0.830632 | 5.282995 | 1.27E-07 |
| ubiquitination | 12 | rs4766428 | 2.61E-17 | 109813508…111837285 | *ANAPC7* | 0.824378 | -0.962298 | 0.3359 |
| synapse | 16 | rs3814883 | 8.82E-15 | 30016830…30034591 | ***DOC2A*** | 0.521296 | 6.088335 | 1.14E-09 |
| synapse | 4 | rs61405217 | 5.39E-11 | 170533784…170644824 | ***CLCN3*** | 0.718835 | 5.771783 | 7.84E-09 |
| synapse | 2 | rs1451488 | 6.72E-17 | 198669426…199437305 | ***PLCL1*** | 0.035058 | 4.910285 | 9.09E-07 |

### SUPPLEMENTAL FIGURES

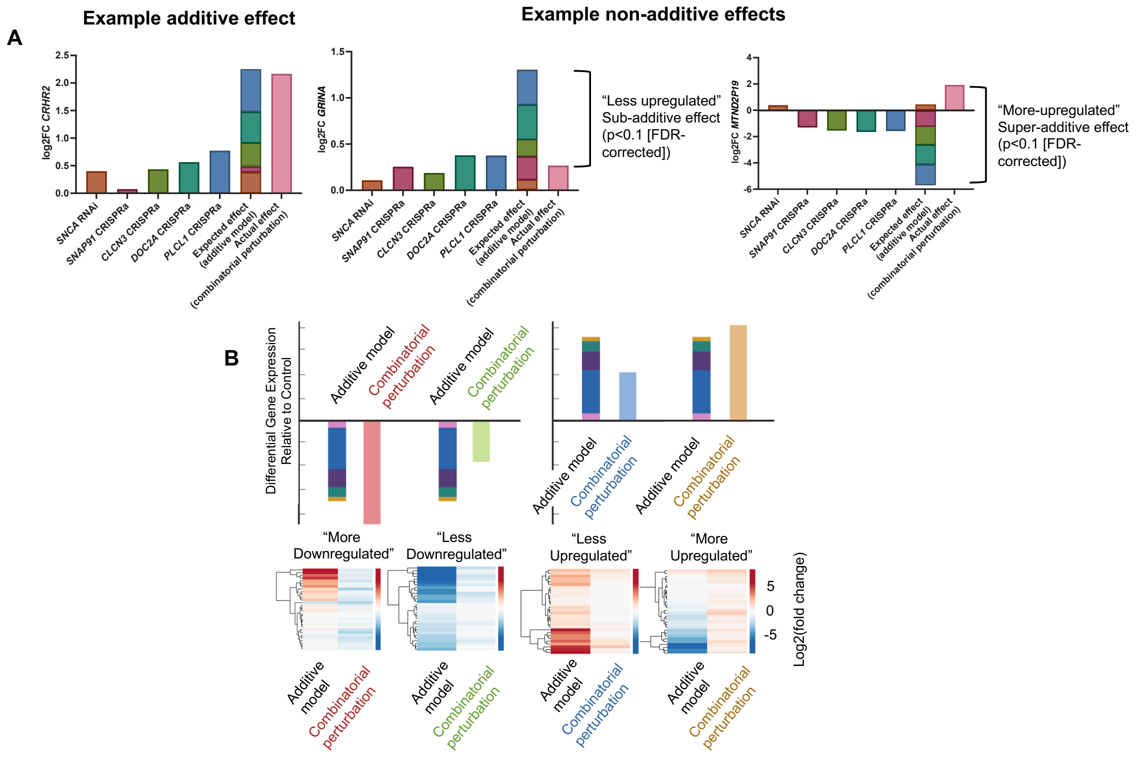


**Box 1: Summary of additive and non-additive effects of joint eGene perturbation on transcription, related to Figure 3, Supplemental Figures 3-5.**

**A.** Example of genes demonstrating additive (*CRHR2*) and non-additive (*GRINA, MTNDP19*) effects following joint perturbation of 5 synaptic eGenes. The summed individual impact of eGene perturbations on *CRHR2* expression did not significantly differ from the impact of joint eGene perturbation, while the impact of joint eGene perturbation on *GRINA* expression was significantly reduced relative to the summed impact of individual eGene perturbations, suggesting saturation or redundancy of individual effects. In contrast to this, the impact of joint eGene perturbation on *MTNDP19* expression was significantly reversed relative to the summed impact of individual eGene perturbations, suggesting antagonistic summation of individual effects. **B.** Example figure showing different categories of non-additive effect found when comparing the impact of combinatorial perturbation relative to that in the predicted additive model. The top row shows exemplar effects on gene expression in the predicted additive model and in the joint perturbation condition. The bottom row shows exemplar heatmaps of differential gene expression for both conditions in each category. "More downregulated" and "more upregulated" categories show more dramatic or opposing effects on gene expression relative to the additive model, while "less downregulated" and "less upregulated" categories show smaller impacts on transcription in the joint perturbation condition when compared to the additive model. In the current study, typically non-additive effects reflected joint eGene perturbation resulting in smaller effects on transcription than the summed individual effects predicted in the additive model ("less downregulated" and "less upregulated" categories).

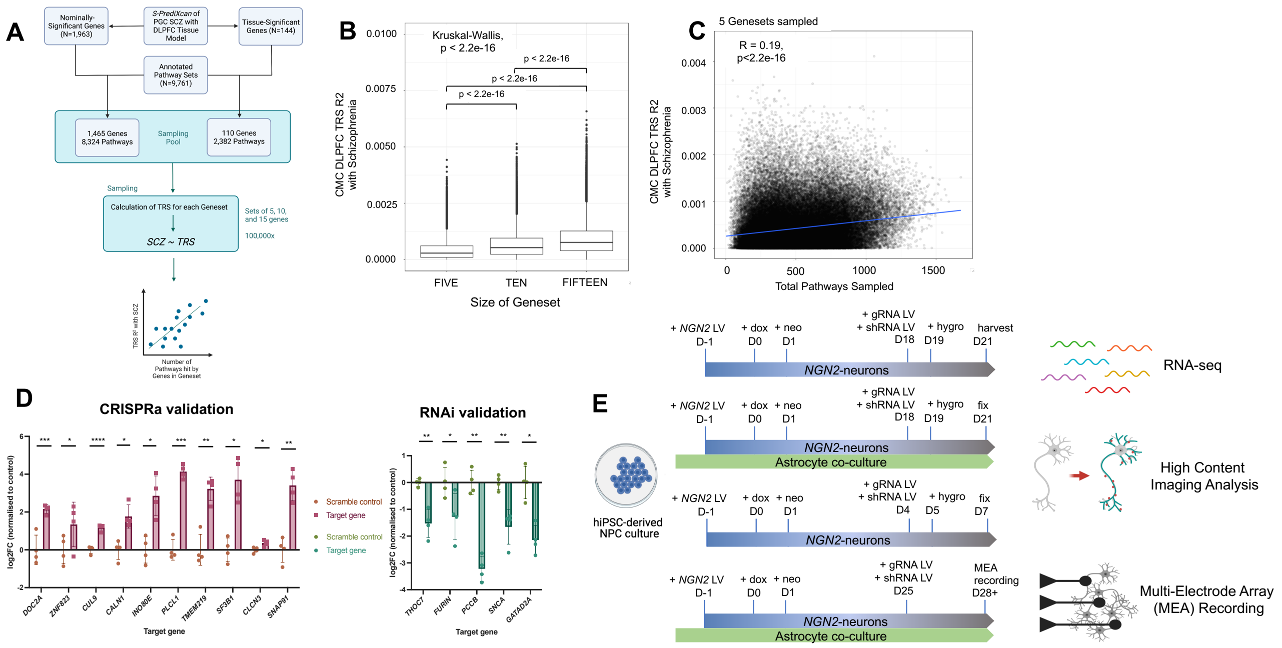


**Supplemental Figure 1. SCZ eGenes impact SCZ case status in a dose-dependent manner and can be perturbed in iGLUTs using CRISPR activation and RNA interference, related to Figure 1.**

**A.** Schematic of random sampling of SCZ gene-sets for TRS. Random sets of 5, 10, and 15 genes were sampled from a pool of either nominally-significant, or tissue-specific-significant SCZ eGenes. TRS were calculated for each gene set, and pathways annotated for the genes included in each randomly sampled gene set. **B.** Boxplot demonstrating increasing TRS $R^2$ with SCZ with increasing number of genes sampled. **C.** Correlation of the number of pathways hit by randomly sampled sets of 5 nominally-significant SCZ genes with the geneset TRS $R^2$ with SCZ. **D.** Validation of CRISPR activation and RNA interference in perturbing target eGenes in D7 hiPSC-NPC derived iGLUTs. 3-5 gRNA or shRNA vectors were tested per target eGene; successful vectors used for subsequent experiments shown in panel. One-way ANOVA with posthoc Dunnett's multiple comparisons test, * = $p<0.05$; ** = $p<0.01$; *** = $p<0.001$; **** = $p<0.0001$. **E.** Schematic showing iGLUT induction, differentiation and perturbation timelines for each phenotyping method: RNA-seq, synapse detection and neurite tracing using high content imaging, and MEA recording. All eGene perturbations were initiated 3 days prior to harvesting, fixing or recording glutamatergic cultures. A minimum of two independent experiments were performed on two healthy donor lines for each phenotyping method.

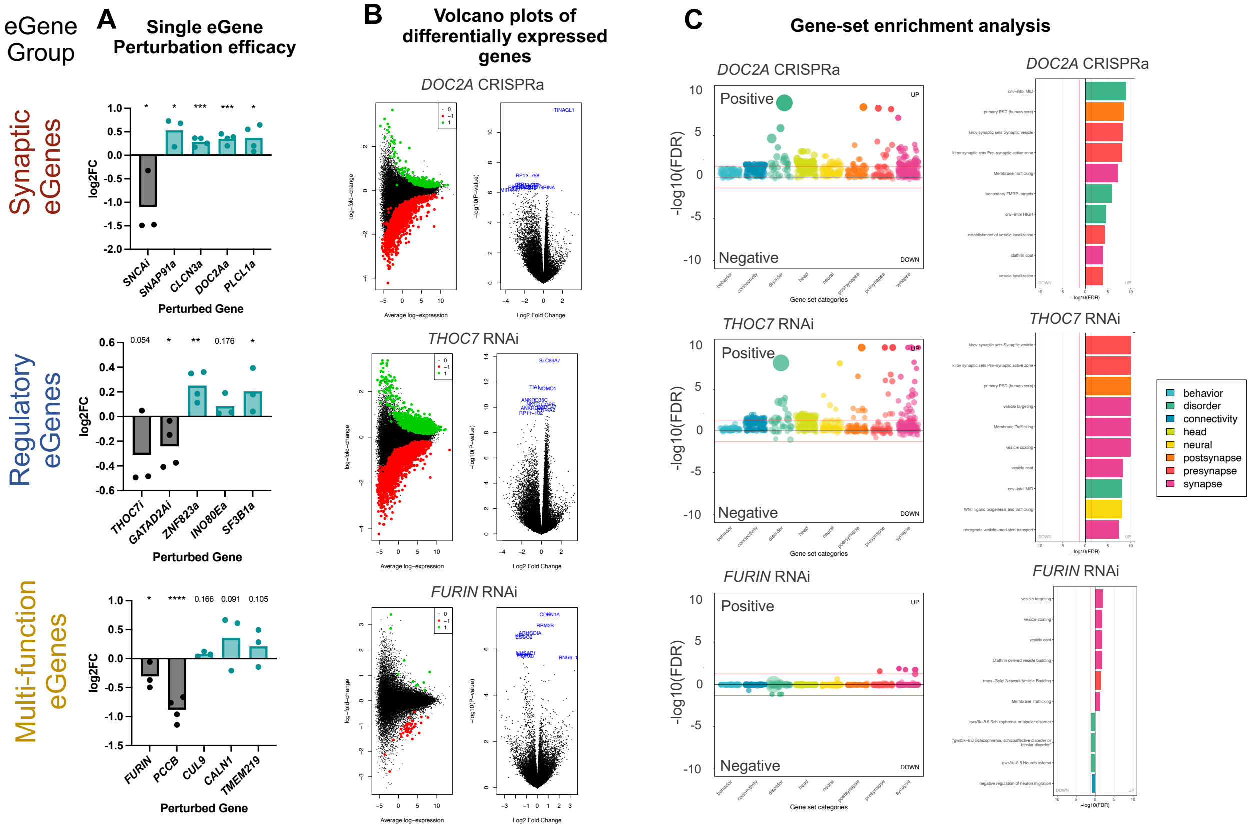


**Supplemental Figure 2. Perturbation of SCZ eGenes results in differential expression of genes relating to brain disorders and synaptic function, related to Figure 2.**

**A.** Log2(fold change) of target eGenes following single perturbations across all three eGene groups in D21 hiPSC-NPC derived iGLUTs. One-tailed t-test, * = $p<0.05$; ** = $p<0.01$; *** = $p<0.001$; **** = $p<0.0001$. **B.** Volcano plots of differential gene expression in perturbation conditions relative to scramble control vectors. Data for one example eGene per eGene group shown: *DOC2A* (Synaptic), *THOC7* (Regulatory) & *FURIN* (Multi-function). **C.** Gene set enrichment analysis (GSEA) performed across a collection of 698 manually curated gene-sets with a neural theme revealed enrichments of genes related to brain disorders (*DOC2A*, *THOC7*) and synaptic functions (*DOC2A*, *THOC7, FURIN*) across multiple eGene perturbations.

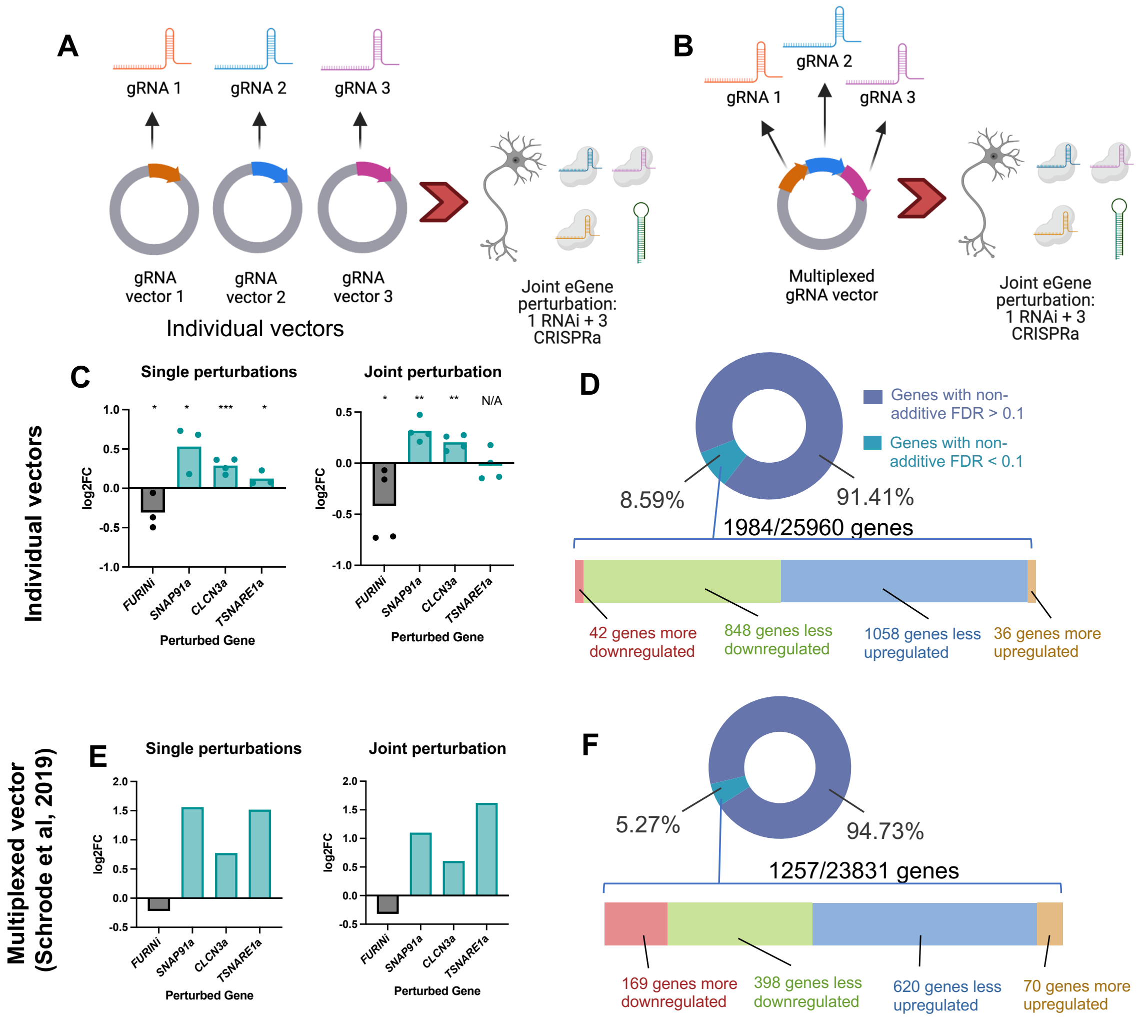


**Supplemental Figure 3: Comparison of non-additive effects of joint perturbations using combined single vectors vs multiplexed vector, related to figure 3.**

**A & B.** Four SCZ eGenes (*FURIN*, *SNAP91*, *CLCN3* and *TSNARE1*) were individually and jointly perturbed using gRNA and shRNA vectors, either using separate (**A**, here) or multiplexed (**B**, previous) vectors in an independent replication of the combinatorial eGene perturbation experiment conducted by Schrode *et al* (2019) using a multiplexed gRNA vector. **C.** Log2(fold change) of four eGenes following single (left) and joint (right) perturbations across in D21 hiPSC-NPC derived iGLUTs, using individual vectors. One-tailed t-test, * = $p<0.05$; ** = $p<0.01$; *** = $p<0.001$; **** = $p<0.0001$. **D.** Summary of non-additive effects of joint eGene perturbation using individual vectors across the transcriptome. 8.59% of genes demonstrated significant non-additive effects; most non-additive effects showed less differential expression following joint perturbation than predicted by the additive model (see **Box 1** for an explanation of different categories of non-additive effects on transcription). **E.** Log2(fold change) of target eGenes following single and joint perturbations across in D21 hiPSC-NPC derived iGLUTs. RNA-seq data taken from Schrode *et al* (2019), using multiplexed vectors. **F.** Summary of non-additive effects of joint eGene perturbation using multiplexed vectors across the transcriptome.

5.27% of genes demonstrated significant non-additive effects. Again, the majority of non-additive effects showed less differential expression following joint perturbation than predicted by the additive model. RNA-seq data taken from Schrode *et al* (2019).

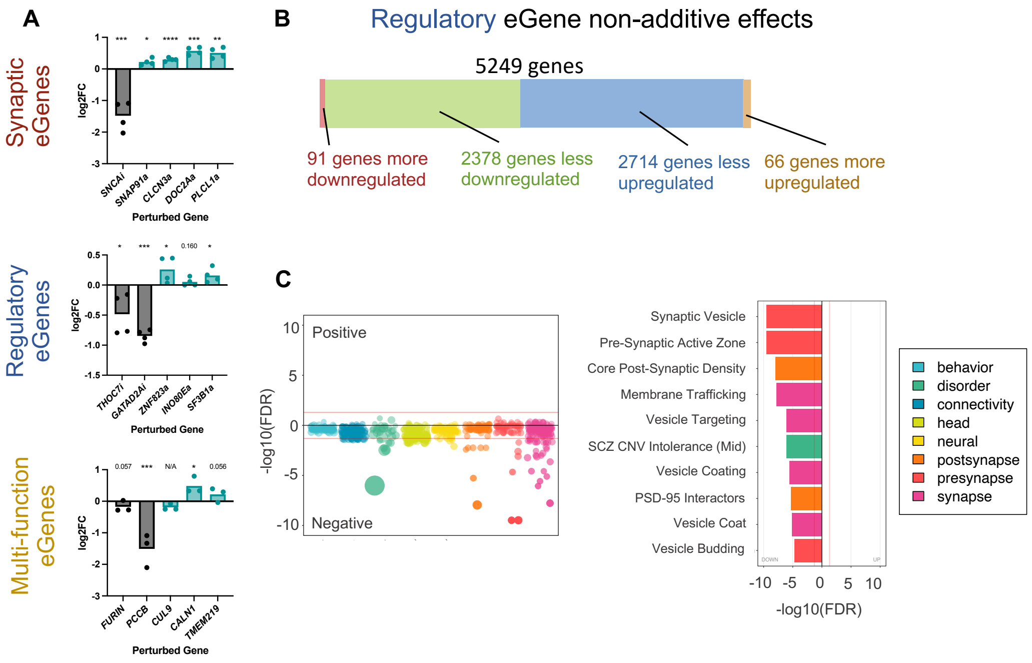


**Supplemental Figure 4. Combinatorial perturbation of regulatory eGenes results in non-additive effects on transcription impacting expression of genes linked to brain disorders and synaptic function, related to Figure 3.**

**A.** Log2(fold change) of target eGenes following joint perturbations across all three eGene groups in D21 hiPSC-NPC derived iGLUTs. One-tailed t-test, * = $p<0.05$; ** = $p<0.01$; *** = $p<0.001$; **** = $p<0.0001$. **B** The majority (>95%) of non-additive genes in the Regulatory eGene set demonstrated significantly less differential expression in the measured combinatorial perturbation relative to the expected additive model (see **Box 1** for an explanation of different categories of non-additive effects on transcription). **C.** GSEA of non-additive genes in the Regulatory eGene set demonstrated significant enrichment for genes relating to brain disorders and synaptic function. SCZ = schizophrenia, CNV = copy number variant, FDR = false discovery rate.

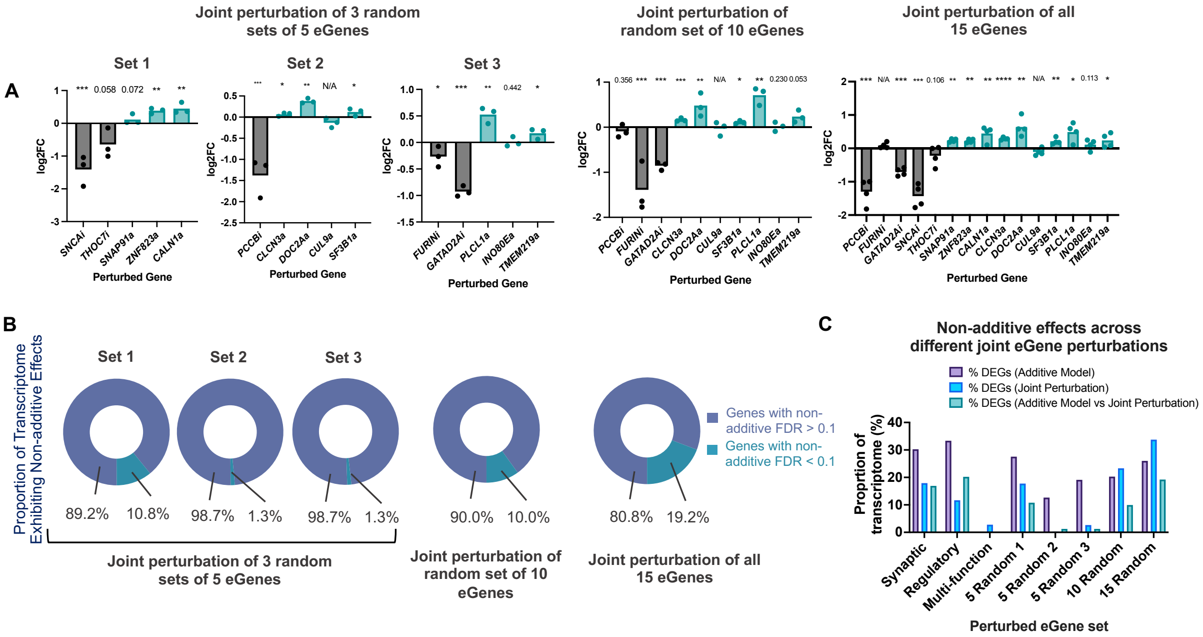


**Supplemental Figure 5. Combinatorial perturbation of random eGenes results in non-additive impacts on transcription which scale with the number of perturbed eGenes, related to Figure 3.**

**A.** Log2(fold change) of target eGenes following combinatorial perturbation of sets of five, ten, and fifteen eGenes randomly assigned from the synaptic, regulatory and multi-function eGene groups. One-tailed t-test, * = p<0.05; ** = p<0.01; *** = p<0.001; **** = p<0.0001. **B.** Non-additive effects across random joint eGene perturbations. The proportion of the transcriptome exhibiting significant non-additive effects increased with increasing numbers of perturbed eGenes (average of 4.5%, 10.0% and 19.2% of the transcriptome with non-additive FDR<0.1 after joint perturbations of five, ten, and fifteen eGenes respectively). **C.** Summary of differentially expressed genes (DEGs) across the additive model, combinatorial perturbation, and the additive-combinatorial comparison for each functional and random eGene set.

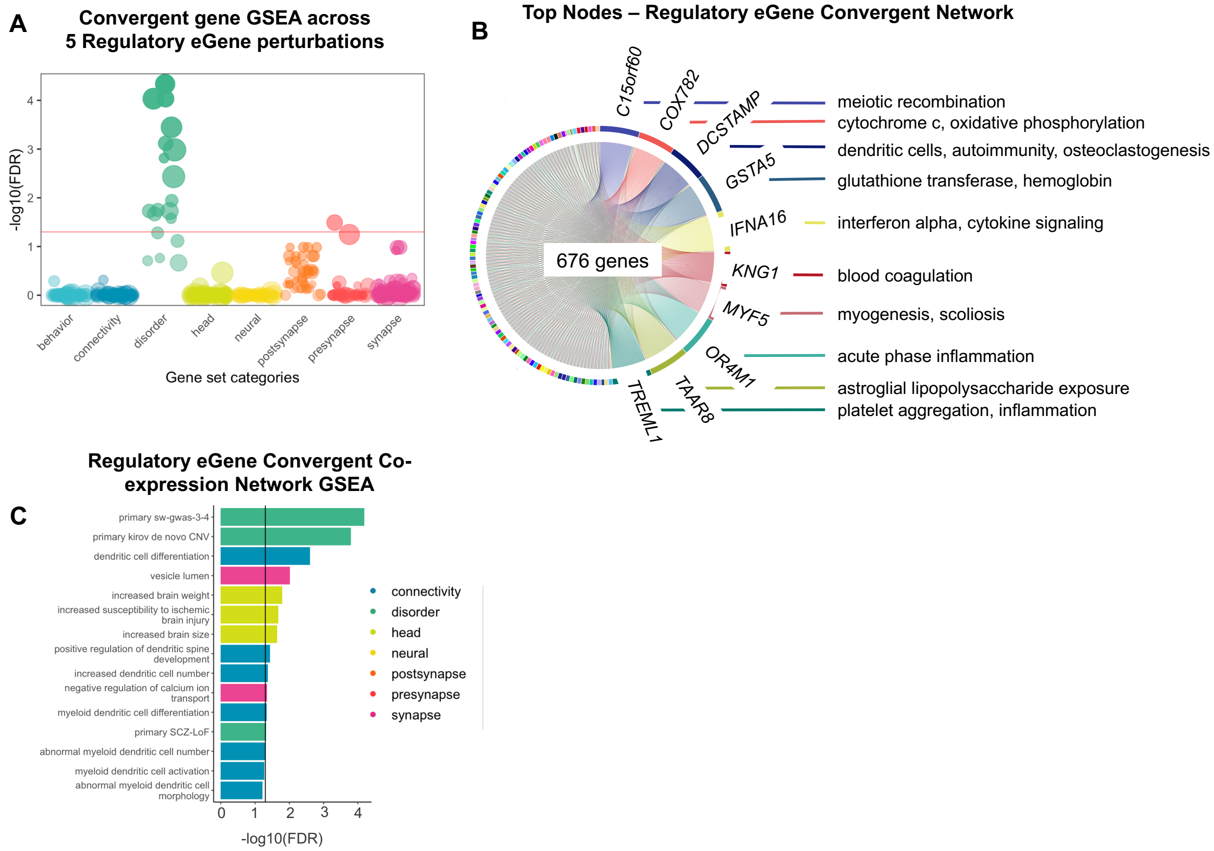


**Supplemental Figure 6. Perturbation of regulatory SCZ eGenes results in transcriptional changes which converge on genes linked to brain disorders and development, related to Figure 4.**

**A.** GSEA of convergent genes in the Regulatory eGene set demonstrated significant enrichment for genes relating to brain disorders. **B.** Bayesian biclustering identified significant convergence of co-expression networks unique to synaptic pathways that replicated in over 12.5% of iterations. Major node genes mediating convergent networks of Regulatory eGenes included *IFNA16*, a cytokine signaling gene, and *OR4M1*, involved in acute inflammatory response. **C.** GSEA of co-expressed network genes in the Regulatory eGene set demonstrated significant enrichment for genes relating to brain development and schizophrenia risk. GSEA = geneset enrichment analysis; SCZ = schizophrenia; GWAS = genome-wide association study; CNV = copy number variant; LoF = loss of function; FDR = false discovery rate.

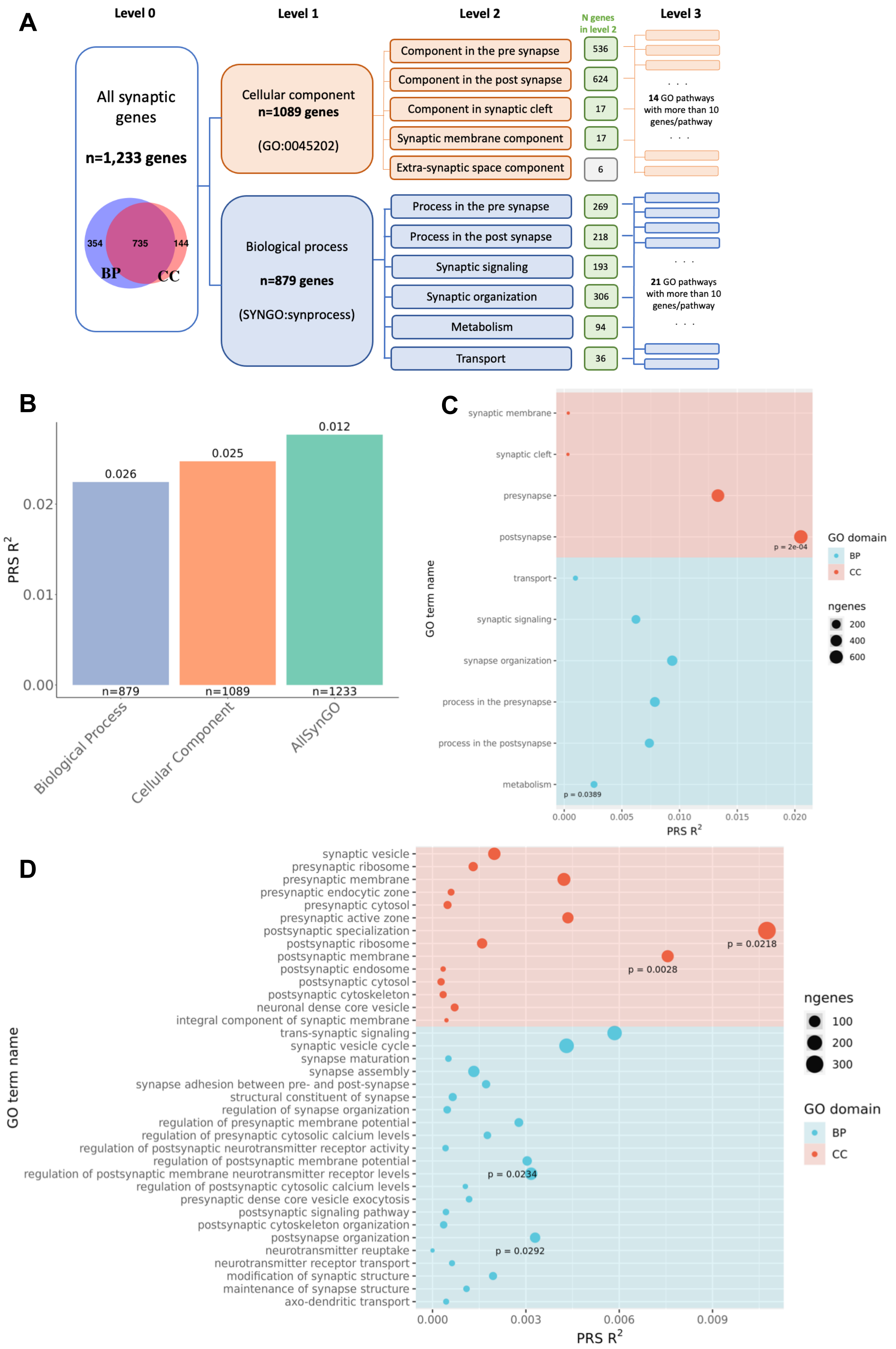
A
Level 0
Level 1
Level 2
N genes in level 2
Level 3
All synaptic genes
n=1,233 genes
354
735
144
BP
CC
Cellular component
n=1089 genes
(GO:0045202)
Component in the pre synapse
536
Component in the post synapse
624
Component in synaptic cleft
17
Synaptic membrane component
17
Extra-synaptic space component
6
14 GO pathways with more than 10 genes/pathway
Biological process
n=879 genes
(SYNGO:synprocess)
Process in the pre synapse
269
Process in the post synapse
218
Synaptic signaling
193
Synaptic organization
306
Metabolism
94
Transport
36
21 GO pathways with more than 10 genes/pathway
B
PRS R2
0.026
0.025
0.012
n=879
n=1089
n=1233
Biological Process
Cellular Component
AllSynGO
C
synaptic membrane
synaptic cleft
presynapse
postsynapse
transport
synaptic signaling
synapse organization
process in the presynapse
process in the postsynapse
metabolism
p = 2e-04
p = 0.0389
GO term name
PRS R2
GO domain
BP
CC
ngenes
200
400
600
D
synaptic vesicle
presynaptic ribosome
presynaptic membrane
presynaptic endocytic zone
presynaptic cytosol
presynaptic active zone
postsynaptic specialization
postsynaptic ribosome
postsynaptic membrane
postsynaptic endosome
postsynaptic cytosol
postsynaptic cytoskeleton
neuronal dense core vesicle
integral component of synaptic membrane
trans-synaptic signaling
synaptic vesicle cycle
synapse maturation
synapse assembly
synapse adhesion between pre- and post-synapse
structural constituent of synapse
regulation of synapse organization
regulation of presynaptic membrane potential
regulation of presynaptic cytosolic calcium levels
regulation of postsynaptic neurotransmitter receptor activity
regulation of postsynaptic membrane potential
regulation of postsynaptic membrane neurotransmitter receptor levels
regulation of postsynaptic cytosolic calcium levels
presynaptic dense core vesicle exocytosis
postsynaptic signaling pathway
postsynaptic cytoskeleton organization
postsynapse organization
neurotransmitter reuptake
neurotransmitter receptor transport
modification of synaptic structure
maintenance of synapse structure
axo-dendritic transport
p = 0.0218
p = 0.0028
p = 0.0234
p = 0.0292
GO term name
PRS R2
ngenes
100
200
300
GO domain
BP
CC

**Supplemental Figure 7. Within-pathway non-additive genes are associated with SCZ risk, related to Figure 5.**

**A.** Diagram depicting the structure of the Synaptic Gene Ontology and number of genes included in each gene set. Each box represents one gene-set used as pathway. **B.** Results for pathway PRS of SYNGO gene-sets (Level 0 and Level 1). Number on top of the bar represents competitive P-value calculated using 10,000 permutations (See Methods). Number on bottom of the bar represents the number of genes included in the pathway. **C**. Results for pathway PRS of SYNGO gene-sets (Level 2). **D.** Results for pathway PRS of SYNGO gene-sets (Level 3). For panels c) and d), p indicated next to the dots correspond to competitive P-value calculated using 10,000 permutations.

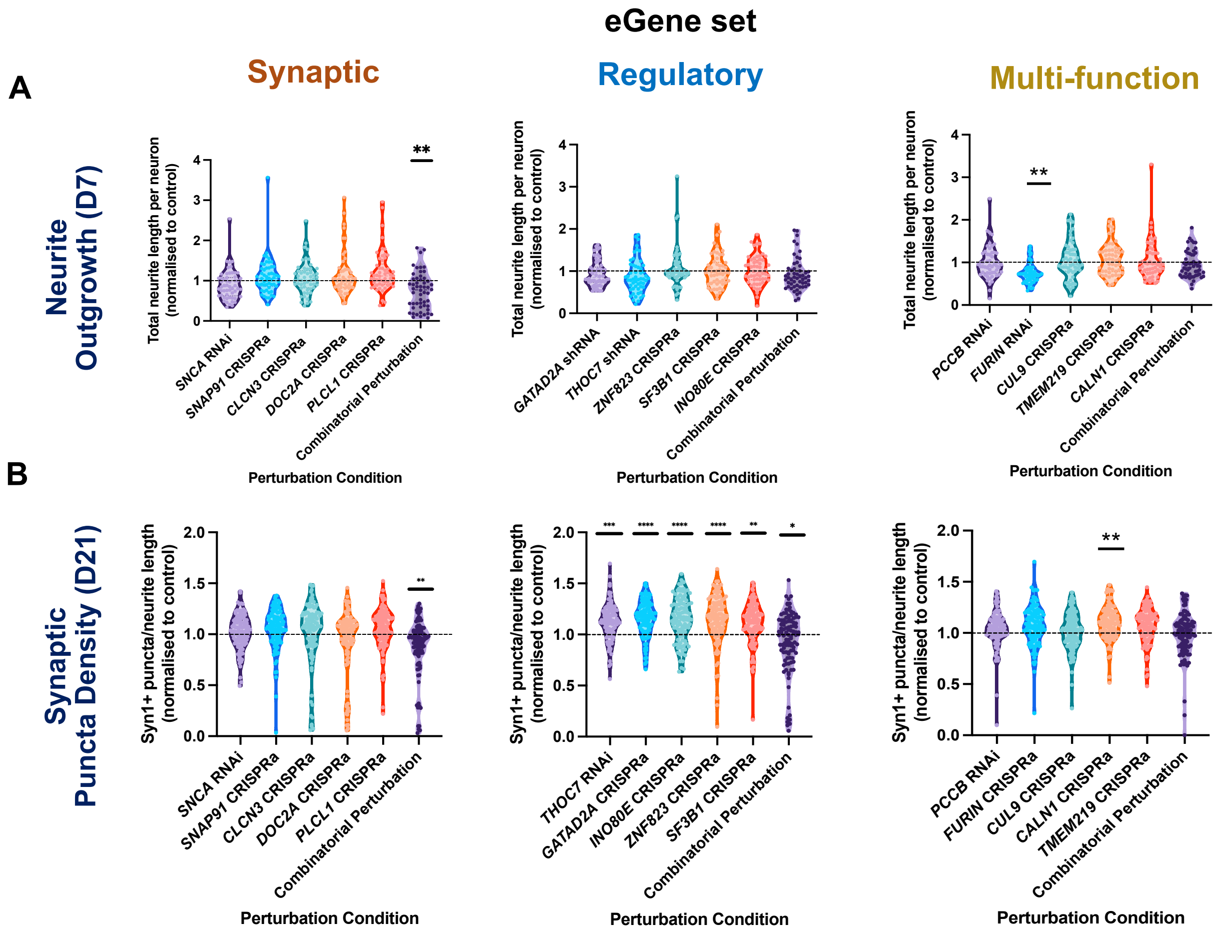


**Supplemental Figure 8. Combinatorial perturbation of SCZ eGenes within, but not across functional categories results in impaired neurite outgrowth and synaptic expression, related to Figure 6.**

**A.** Impact of single and joint eGene perturbations on early (D7) neurite outgrowth in hiPSC-derived iGLUTs across all three eGene groups. Combinatorial perturbation of all five synaptic eGenes and individual perturbation of *FURIN* (Multi-function eGene set) resulted in significant decreases in early neurite outgrowth relative to scramble control conditions. **B.** Impact of single and joint eGene perturbations on Synapsin1 (SYN1)-positive puncta expression in D21 hiPSC-derived iGLUTs across all three eGene groups. SYN1+ puncta values are expressed relative to MAP2-positive neurite length in each image. Combinatorial perturbation of all five synaptic eGenes and all five regulatory eGenes resulted in significant decreases in synaptic puncta expression relative to scramble control conditions. Individual perturbation of regulatory eGenes and *CALN1* (Multi-function eGene set) resulted in significant increases of SYN1+ puncta density relative to scramble controls. N = minimum of 2 independent experiments across 2 donor lines with 12 technical replicates per condition and 9 images analysed per replicate. One-way ANOVA with post-hoc Bonferroni multiple comparisons test. * = p<0.05; ** = p<0.01; *** = p<0.001; **** = p<0.0001.

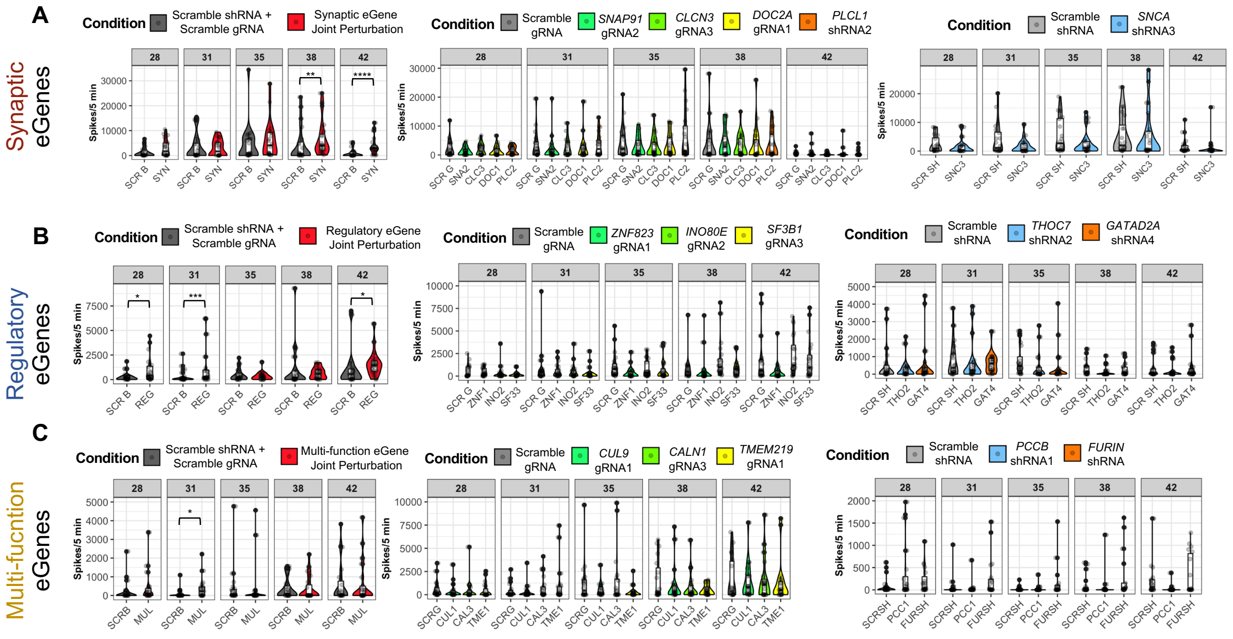


**Supplemental Figure 9. Combinatorial perturbation of SCZ eGenes within, but not across functional categories results in neuronal hyperactivity, related to Figure 6.**

**A-C.** Impact of single and joint eGene perturbations on MEA-recorded spike traces in mature (4-6 week-old) hiPSC-derived iGLUT/astrocyte co-cultures across all three eGene groups. **A.** Combinatorial perturbation of all five synaptic eGenes resulted in transient neuronal hyperactivity relative to a scramble control condition. **B.** Combinatorial perturbation of all five regulatory eGenes resulted in transient neuronal hyperactivity relative to a scramble control condition. **C.** Combinatorial perturbation of all five multi-function eGenes did not result in the same pattern of transient neuronal hyperactivity seen in the synaptic and regulatory combinatorial perturbations. Individual eGene perturbations did not significantly impact neuronal firing frequency. N = minimum of 2 independent experiments across 2 donor lines with 20 technical replicates per condition. One-way ANOVA with post-hoc Bonferroni multiple comparisons test. * = $p<0.05$; ** = $p<0.01$; *** = $p<0.001$; **** = $p<0.0001$.
